# Comparison of low eGFR prevalence and prediction for mortality using 2009 and 2021 CKD-EPI equations in Mexican adults

**DOI:** 10.1101/2025.05.23.25328234

**Authors:** Daniel Ramírez-García, Carlos A. Fermín-Martínez, Paulina Sánchez Castro, Brenda Guadalupe Cortez-Flores, Jerónimo Perezalonso Espinosa, Juan Pablo Díaz-Sánchez, Karime Berenice Carrillo-Herrera, Leslie Alitzel Cabrera-Quintana, Jaime Berumen-Campos, Pablo Kuri-Morales, Roberto Tapia-Conyer, Jacqueline A. Seiglie, Jesus Alegre-Díaz, Neftali Eduardo Antonio-Villa, Omar Yaxmehen Bello-Chavolla

## Abstract

Accurate estimation of glomerular filtration rate (eGFR) is essential for identifying and managing chronic kidney disease (CKD). The CKD-EPI 2021 equation removed the race coefficient from the 2009 version, but its impact in Mexican populations remains unclear. Here, we compared eGFR category prevalence, and predictive performance between the CKD-EPI 2009 and 2021 creatinine-based eGFR equations, as well as the prognostic relevance of reclassification in eGFR categories using the 2021 equation in Mexicans. We evaluated 25,110 adults _≥_20 years from the 2016-2023 cycles of the Mexican National Health and Nutrition Survey (ENSANUT) to estimate national low eGFR and eGFR category prevalence using both equations. We also assessed 5-year and 10-year risk of all-cause, cardiovascular, and kidney-related mortality in 142,884 adults from the Mexico City Prospective Study (MCPS) using Cox proportional hazards and Fine & Gray regression models. In ENSANUT 2023, prevalence of eGFR <60mL/min/1.73m2 was lower with CKD-EPI 2021 (2.9%, 95%CI 1.56–4.24%) compared to the 2009 equation (3.6%, 95%CI 1.99–5.21). Use of the 2021 equation resulted in upward eGFR reclassification in 6.52% (95%CI 4.07-8.97) of adults _≥_20 years, particularly among older adults and those with hypertension or diabetes, yielding a reduction in 486,532 adults identified with eGFR <60mL/min/1.73m^2^ compared to the 2009 equation. In MCPS, despite both equations showing similar C-statistics, the 2021 equation showed slightly improved predictive performance for 5-year and 10-year mortality outcomes. The 2021 equation reclassified 8.3% of participants to higher eGFR categories, and reclassification was associated with decreased risk of all-cause, cardiovascular, and kidney-related mortality, particularly for participants reclassified upward from G3a-G5 categories. The CKD-EPI 2021 equation yields lower prevalence of low eGFR but leads to prognostically relevant eGFR category reclassification compared to the 2009 equation. Our findings support the implementation of the 2021 equation for population health monitoring in Mexico without compromising prognostic utility.

## INTRODUCTION

Chronic kidney disease (CKD) has become a significant health problem worldwide, especially in low- and middle-income countries, where inequities in access to preventive services, renal replacement therapy (RRT), and the increasing prevalence of cardio-metabolic diseases have contributed to growing prevalence and mortality rates^1^. CKD diagnosis and stratification typically involves estimation of the glomerular filtration rate (eGFR) and measuring albuminuria^2^. Previous clinical guidelines recommended using the 2009 CKD Epidemiology Collaboration (CKD-EPI) equation which accounts for age, sex, and race-related variability^3,4^. However, using race in clinical algorithms has been recently questioned, considering it as a social and not a biological construct^5^. Consequently, a new CKD-EPI equation without a race coefficient was developed in 2021^6^; which, compared to directly measured GFR, leads to higher CKD prevalence estimates amongst black individuals, and lower amongst non-black individuals^6^.

Mexicans have an admixed ancestry, comprising of ∼50% of Amerindian native-American, 45% European and <5% African and East Asian contributions^7^. This admixture profile contrasts with the predominantly European ancestry of non-Hispanic white individuals, and African ancestry of non-Hispanic Black individuals in the US^8^. Despite representing a large share of the US population, Mexicans and broader Latino populations remain underrepresented in kidney disease studies, despite showing significant inter-population variability in CKD markers^9,10^. Furthermore, eGFR equations commonly used in clinical settings frequently classify Mexican individuals as white, disregarding emerging evidence of population-specific differences in serum creatinine levels^10^, and the influence of genetic ancestry on the accuracy of creatinine-based eGFR estimations^11^. To date, no direct estimations of CKD prevalence have been conducted in Mexico, with prior studies focusing on forecasting estimates based on statistical models^1^. Moreover, the impact of replacing the 2009 with the 2021 equation on CKD prevalence, and the utility of these equations for prediction of relevant outcomes, including all-cause and cause-specific mortality in Mexican populations, has not been previously validated. In this study, we aimed to estimate the prevalence of eGFR categories and low eGFR using the 2009 and 2021 CKD-EPI equations in adults using nationally representative surveys from 2016 to 2023 in Mexico, and to assess the performance of both equations for prediction of all-cause, cardiovascular, and kidney-related mortality in prospective cohort of adults living in Mexico City.

## METHODS

### Study populations

#### Mexican National Health and Nutrition Survey

We used data from the cross-sectional Mexican National Health and Nutrition Survey (ENSANUT) cycles 2016, 2018, 2020, 2021, 2022, and 2023. ENSANUT is a probabilistic survey representative at a regional and national level which uses a multi-stage probabilistic cluster stratified sampling design^12^. Sample weights are corrected using the response rate observed from the immediate previous cycle and calibrated to account for representation of specific subgroups. Participants completed a questionnaire including demographic, socioeconomic, and health-related information, along with a physical examination. Additionally, serum samples were collected from a random subsample per cycle to measure fasting glucose, serum creatinine, lipid profile, and glycated hemoglobin^12^. Further details on ENSANUT’s methodology are reported elsewhere^12–15^, and in **Supplementary Methods**. For this study, we included individuals _≥_20 years with measured serum creatinine to calculate eGFR.

#### Mexico City Prospective Study

The Mexico City Prospective Study (MCPS) is a population-based, prospective cohort study with a baseline survey conducted from 1998 to 2004 on two urban districts of Mexico City^16^. In total, 159,755 individuals aged ≥35 years were recruited and underwent a comprehensive questionnaire collecting sociodemographic and health-related information, clinical measurements (anthropometry, and blood pressure), and a non-fasting 10-mL blood sample. We included all participants with complete mortality, covariate, and serum creatinine data. Mortality follow-up of participants was done through electronic probabilistic linkage to death registries, with deaths tracked up to September 30^th^, 2022, and registered according to the International Classification of Diseases 10th Revision (ICD-10)^17^. Mortality was codified in accordance with the underlying causes of death determined by study clinicians as all-cause, cardiovascular disease (CVD) (myocardial infarction for codes I20-I25, stroke for codes I60-I69), and kidney-related mortality (codes N17-N19, E10-E14, I12-I13, N28, Q61, N00-N16, Y84). Details on approval, mortality follow-up, comorbidities, and creatinine measurements are reported in **Supplementary Materials**.

#### Estimated glomerular filtration rate and chronic kidney disease

eGFR was calculated using the CKD-EPI 2009 and 2021 creatinine-based equations^6^, and classified into six categories as recommended by KDIGO guidelines, specifically: G1 normal or high (_≥_90 mL/min/1.73m^2^), G2 mildly decreased (60–89 mL/min/1.73m^2^), G3a mildly to moderately decreased (45–59 mL/min/1.73m^2^), G3b moderately-to-severely decreased (30–44 mL/min/1.73 m^2^), G4 severely decreased (15-29 mL/min/1.73m^2^), and G5 kidney failure (<15 mL/min/1.73m^2^)^3^. Given that albuminuria was not measured in either study, low eGFR was used as a proxy for CKD, defined as eGFR <60 mL/min/1.73m^2^. We also explored the prevalence of CKD defined as the combination of previous self-reported medical diagnosis of CKD or low eGFR.

### Statistical analyses

We obtained weighted prevalence estimates of eGFR categories and low eGFR using the 2021 CKD-EPI creatinine-based equation, considering complex survey design and sample weights from the ENSANUT venous blood subsample using the *survey* R package. Additionally, we estimated low eGFR prevalence stratified by age categories (<60 or ≥60 years old), sex, diabetes, hypertension, and obesity status. Next, we compared these estimations against the 2009 CKD-EPI creatinine-based equation, exploring the proportion, and total number of individuals that would be reclassified to different eGFR categories. Additional details on ENSANUT survey design are available in **Supplementary Methods**.

To evaluate the predictive performance of both CKD-EPI equations for all-cause, CVD, and kidney-related mortality, we fitted a baseline Cox proportional hazards model stratified by baseline age category and adjusted for sex, place of residence, education, smoking, diabetes, history of CVD, systolic blood pressure, and total cholesterol, which is similar to the CKD Prognosis Consortium model for all-cause and CVD mortality risk^18^. eGFR estimated with either the 2009 or 2021 equations, was added separately to this baseline model to assess discrimination and performance metrics^19,20^. For CVD and kidney-related mortality, we used Fine & Gray competing risk models, adjusted for the same covariates using the *survival* R package. Administrative censoring was applied at both 5 and 10 years to assess all outcomes. The non-linear relationship between eGFR and mortality risk was investigated with penalized smoothing splines^21^, and eGFR was modeled using restricted cubic splines with 5 knots^22^. Fixed-time point, and time-range discrimination was assessed using Harrell’s c-statistic, and overall performance was determined evaluating changes in the model’s chi-square statistic, the Brier score, and the scaled Brier score. While relative hazards might not capture a marker’s added predictive value^19^, we report the hazard ratios (HR) and sub-distribution HRs (sHR)s for eGFR 2009 or 2021 from extended models. To assess the prognostic significance of reclassification from eGFR categories based on the 2021 equation to those estimated with the 2009 equation, we generated an indicator variable identifying participants who were 1) reclassified upward from G2 to G1 or 2) reclassified upward from G3a-G5. We then estimated the relative risks for all-cause, cardiovascular, and kidney-related mortality by comparing these participants with those who remained in the same categories (i.e., not reclassified). Outcomes were visualized using Kaplan-Meier curves adjusted using inverse probability of treatment weighting (IPTW) groups with similar covariates as previous models using the *adjustedCurves* R package. Additional details on modeling, missing data, and underlying assumptions are reported in **Supplementary Methods**. All statistical analyses were conducted using R software version 4.4.2.

## RESULTS

### eGFR categories and CKD prevalence in Mexican population

We analyzed data from 25,110 adults _≥_20 years from six ENSANUT cycles during the 2016-2023 period (**Supplementary Table 1**). Using the 2021 CKD-EPI equation, the prevalence of eGFR categories remained stable over time. The proportion of individuals on the G1 category reached 87.08% (95%CI 84.31–89.84%) in 2023, with a slight decrease over time (**Figure 1A**). G2 prevalence was much lower, reaching 9.98% (95%CI 7.76– 12.20%) by 2023 (**Figure 1B**). G3a and G3b categories had a prevalence of 1.76% (95%CI 0.77–2.76%) and 0.90% (95%CI 0.22–1.57%) in 2023, respectively (**Figure 1C**), and G4 and G5 prevalence estimations from 2016 to 2023 were below 0.6% and 0.35%, respectively. (**Table 1**). Categories G3a-G5 were stable over time but presented an increase during the 2020-2023 period. In 2023, low eGFR prevalence was 2.94% (95%CI 1.61–4.28%), remaining stable from 2016 to 2022, and showing a slight increase in 2023 (**Figure 1D**, **Table 1**). Prevalence was similar between men and women, but higher among adults _≥_60 years (**Figure 1E, F**), in individuals with diabetes or hypertension (**Figure 1G, H**), and no differences were observed in individuals with obesity (**Figure 1I**). The estimated number of individuals with low eGFR in Mexico ranged from 1,264,500 (95%CI 822,068– 1,706,931) in 2016 to 2,251,125 (95%CI 1,240,552–3,261,697) in 2023 (**Supplementary Table 2**).

**Figure 1.**
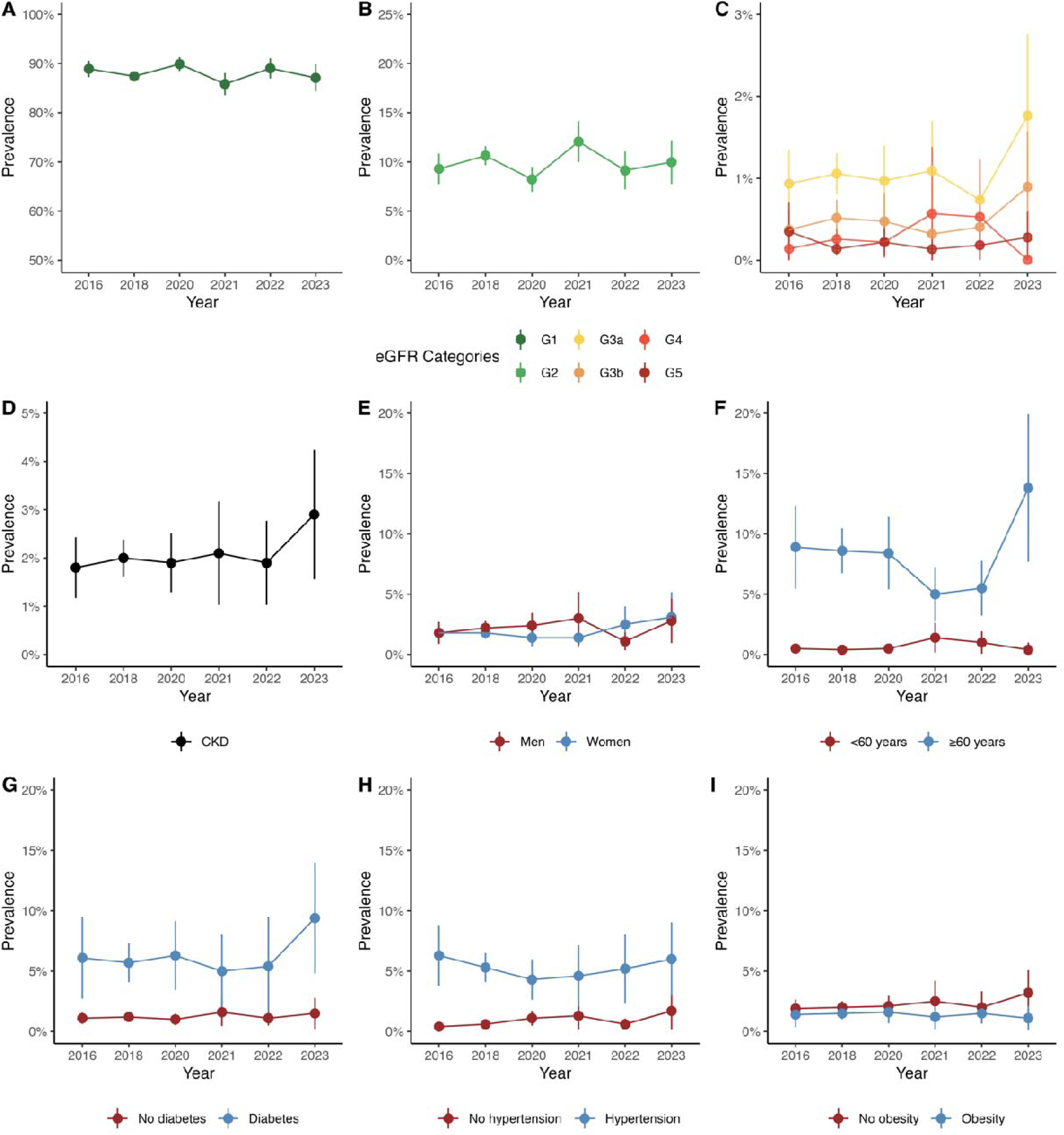
Prevalence of eGFR categories and CKD based on low eGFR in Mexico using the 2021 CKD-EPI creatinine equation in ENSANUT 2016-2023 data. (A) G1, (B) G2, (C) G3a, G3b, G4, and G5, (D) Overall CKD prevalence and CKD prevalence stratified by (E) sex, (F) age category, (G) diabetes, (H) hypertension, and (I) obesity status. G1 normal or high (_≥_90 mL/min per 1.73 m^2^), G2 mildly decreased (60–89 mL/min/1.73 m^2^), G3a mildly to moderately decreased (45–59 mL/min/1.73 m^2^), G3b moderately to severely decreased (30–44 mL/min/1.73 m^2^), G4 severely decreased (15-29 mL/min/1.73 m^2^), and G5 kidney failure (<15 mL/min/1.73 m^2^). eGFR: Estimated glomerular filtration rate, CKD: Chronic Kidney Disease, CKD-EPI: Chronic Kidney Disease Epidemiology Collaboration.

**Table 1.**
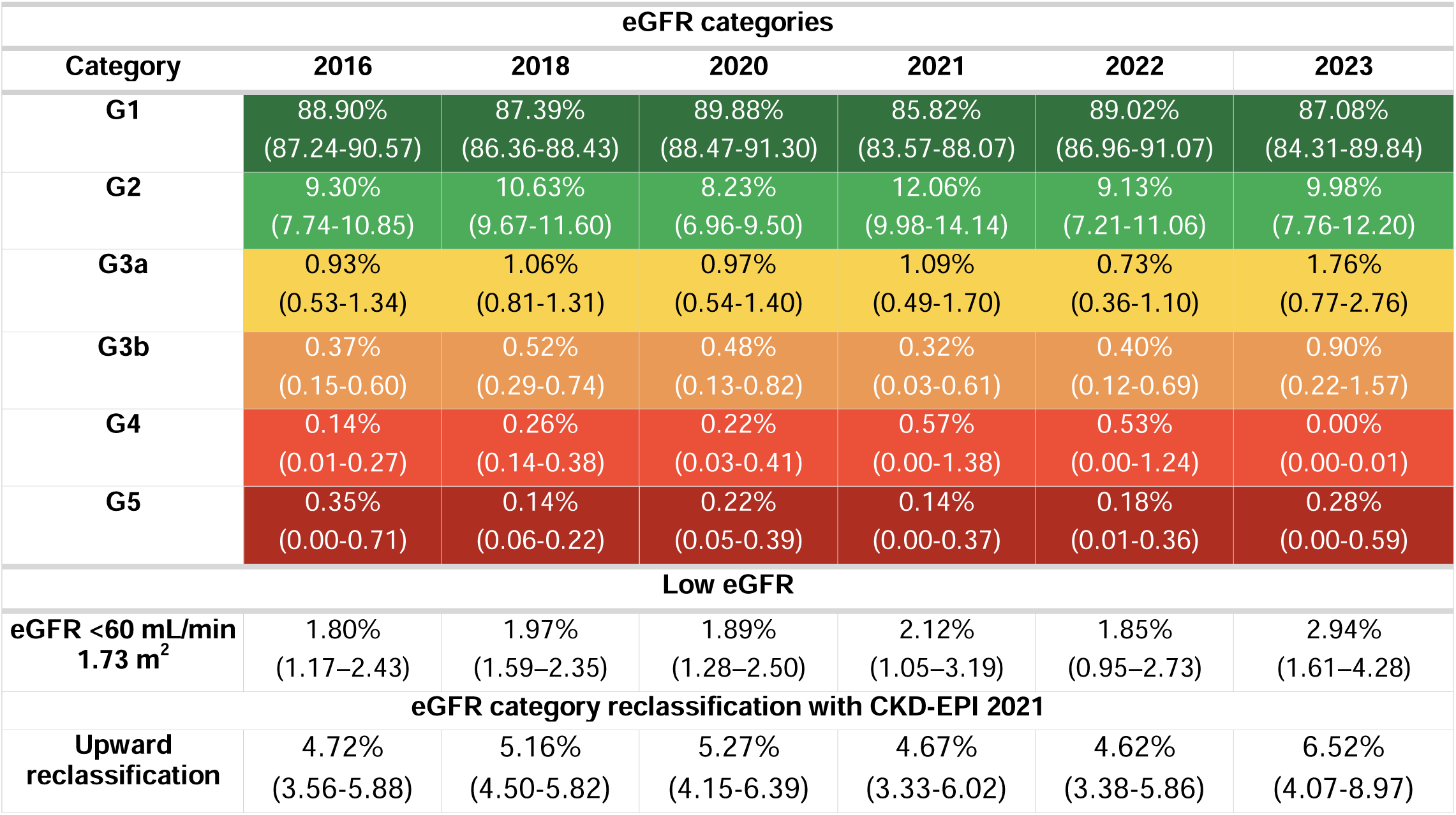
Nationally representative prevalence with 95% confidence intervals from 2016 to 2023 of eGFR categories, low eGFR (<60 mL/min 1.73 m^2^) and eGFR category reclassification using ENSANUT data. eGFR was estimated using the CKD-EPI 2021 creatinine-based equation. G1 normal or high (_≥_90 mL/min per 1.73 m^2^), G2 mildly decreased (60–89 mL/min/1.73 m^2^), G3a mildly to moderately decreased (45–59 mL/min/1.73 m^2^), G3b moderately to severely decreased (30–44 mL/min/1.73 m^2^), G4 severely decreased (15-29 mL/min/1.73 m^2^), and G5 kidney failure (<15 mL/min/1.73 m^2^). The table also shows the prevalence of eGFR category reclassification using the 2021 CKD-EPI equation from categories classified originally using the 2009 CKD-EPI equation. Abbreviations: eGFR: Estimated glomerular filtration rate, CKD-EPI: Chronic Kidney Disease Epidemiology Collaboration.

| eGFR categories |  |  |  |  |  |  |
| --- | --- | --- | --- | --- | --- | --- |
| Category | 2016 | 2018 | 2020 | 2021 | 2022 | 2023 |
| **G1** | 88.90%<br>(87.24-90.57) | 87.39%<br>(86.36-88.43) | 89.88%<br>(88.47-91.30) | 85.82%<br>(83.57-88.07) | 89.02%<br>(86.96-91.07) | 87.08%<br>(84.31-89.84) |
| **G2** | 9.30%<br>(7.74-10.85) | 10.63%<br>(9.67-11.60) | 8.23%<br>(6.96-9.50) | 12.06%<br>(9.98-14.14) | 9.13%<br>(7.21-11.06) | 9.98%<br>(7.76-12.20) |
| **G3a** | 0.93%<br>(0.53-1.34) | 1.06%<br>(0.81-1.31) | 0.97%<br>(0.54-1.40) | 1.09%<br>(0.49-1.70) | 0.73%<br>(0.36-1.10) | 1.76%<br>(0.77-2.76) |
| **G3b** | 0.37%<br>(0.15-0.60) | 0.52%<br>(0.29-0.74) | 0.48%<br>(0.13-0.82) | 0.32%<br>(0.03-0.61) | 0.40%<br>(0.12-0.69) | 0.90%<br>(0.22-1.57) |
| **G4** | 0.14%<br>(0.01-0.27) | 0.26%<br>(0.14-0.38) | 0.22%<br>(0.03-0.41) | 0.57%<br>(0.00-1.38) | 0.53%<br>(0.00-1.24) | 0.00%<br>(0.00-0.01) |
| **G5** | 0.35%<br>(0.00-0.71) | 0.14%<br>(0.06-0.22) | 0.22%<br>(0.05-0.39) | 0.14%<br>(0.00-0.37) | 0.18%<br>(0.01-0.36) | 0.28%<br>(0.00-0.59) |
| Low eGFR |  |  |  |  |  |  |
| **eGFR <60 mL/min**<br>**1.73 m<sup>2</sup>** | 1.80%<br>(1.17-2.43) | 1.97%<br>(1.59-2.35) | 1.89%<br>(1.28-2.50) | 2.12%<br>(1.05-3.19) | 1.85%<br>(0.95-2.73) | 2.94%<br>(1.61-4.28) |
| eGFR category reclassification with CKD-EPI 2021 |  |  |  |  |  |  |
| **Upward**<br>**reclassification** | 4.72%<br>(3.56-5.88) | 5.16%<br>(4.50-5.82) | 5.27%<br>(4.15-6.39) | 4.67%<br>(3.33-6.02) | 4.62%<br>(3.38-5.86) | 6.52%<br>(4.07-8.97) |

### Comparison of low eGFR prevalence using different CKD-EPI equations in ENSANUT

eGFR was consistently higher using the 2021 CKD-EPI equation, with a median of 111.27 (IQR 99.1–121.1) compared to a median of 107.9 (IQR 95.1–119.4) using the 2009 CKD-EPI equation (**Figure 2A**). Consequently, the prevalence of G3a-G5 categories was also higher when using the 2009 compared with the 2021 equation (**Figure 2B, Supplementary Table 3 and 4**). This trend persisted regardless of survey cycle, sex, age, diabetes, or hypertension status (**Figure 2C–F**). The difference between equations was much lower among individuals with higher eGFR (i.e. participants <60 years, without diabetes, or without hypertension). Prevalence estimates of a CKD proxy comprising previous self-reported medical diagnosis of CKD or low eGFR for both equations can be found in **Supplementary Table 5,** and **Supplementary Figure 1**.

**Figure 2.**
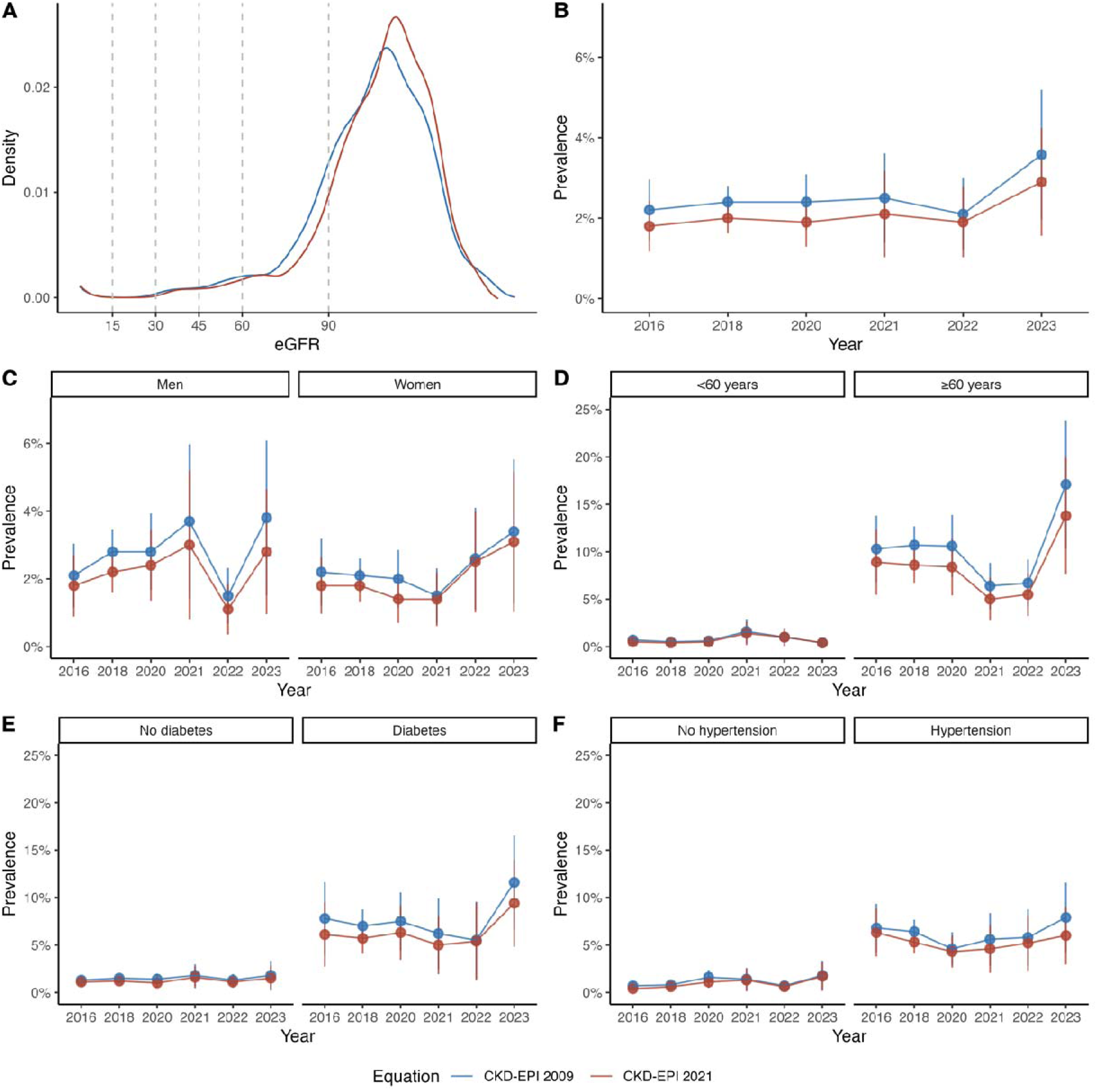
eGFR distribution and comparison of CKD prevalence using either 2021 or 2009 CKD-EPI creatinine equations in ENSANUT 2016-2023 data. (A) eGFR distribution, (B) Comparison of overall CKD prevalence using both equations, (C) Comparison of CKD prevalence in men, (D) age category, (E) diabetes status, and (F) hypertension status. eGFR: Estimated glomerular filtration rate, CKD: Chronic Kidney Disease, CKD-EPI: Chronic Kidney Disease Epidemiology Collaboration.

### Reclassification in eGFR categories using the 2021 CKD-EPI equation

Using the 2021 CKD-EPI equation resulted in reclassification of 6.52% (95%CI 4.07-8.97) of adults to higher eGFR categories in 2023, with similar estimates over the 2016-2023 period (**Table 1**). Reclassified individuals were predominately men, older, had a higher proportion of diabetes, and hypertension compared to those not reclassified (**Supplementary Table 6**). Adoption of 2021 CKD-EPI equation would result in 247,962 less individuals with low eGFR in 2016 and 486,532 less in 2023 (**Supplementary Table 2**); moreover, a total of 4,986,795 individuals (95%CI 3,009,252 to 6,964,338) would be reclassified upwards when using the 2021 CKD-EPI equation in 2023.

### Predictive performance of 2009 and 2021 CKD-EPI equations for cause-specific mortality

Among 159,517 participants recruited from 1998 to 2004 in MCPS, 142,884 had complete covariate and mortality data, and were included in the analyses (**Supplementary Figure 2, Supplementary Table 7**). At the 10-year administrative censoring, there were 12,823 all-cause, 4,277 cardiovascular, and 1,562 kidney-related deaths (**Supplementary Figure 3**). Similar to our findings in ENSANUT, eGFR distribution was higher using the 2021 compared to the 2009 equation (**Figure 3A**), and the prevalence of each eGFR category and low eGFR were lower with the 2021 equation (**Supplementary Tables 8-9**). Risk for all-cause mortality was similar between equations in both extended models, with slightly lower HRs when using the 2021 equation, particularly at higher eGFR values (**Figure 3B**). When evaluating discrimination and overall performance for all-cause, cardiovascular, and kidney-related mortality at 5 and 10-years, we found similar discrimination and performance metrics when adding eGFR calculated with either the 2009 or the 2021 equation to the baseline model. Albeit small in absolute terms, relative larger improvements were observed in Brier scores, Chi-squared statistics and log-likelihood when adding the 2021 equation to the baseline model for all 5-year outcomes and for 10-year cardiovascular mortality. In contrast, adding the 2009 equation to the baseline model improved prediction for 10-year all-cause, and kidney-related mortality **(Figures 3C-D**, **Table 3** and **Supplementary Table 10**). Full report of all fitted Cox and Fine & Gray models are reported in **Supplementary Tables 11-22**.

**Figure 3.**
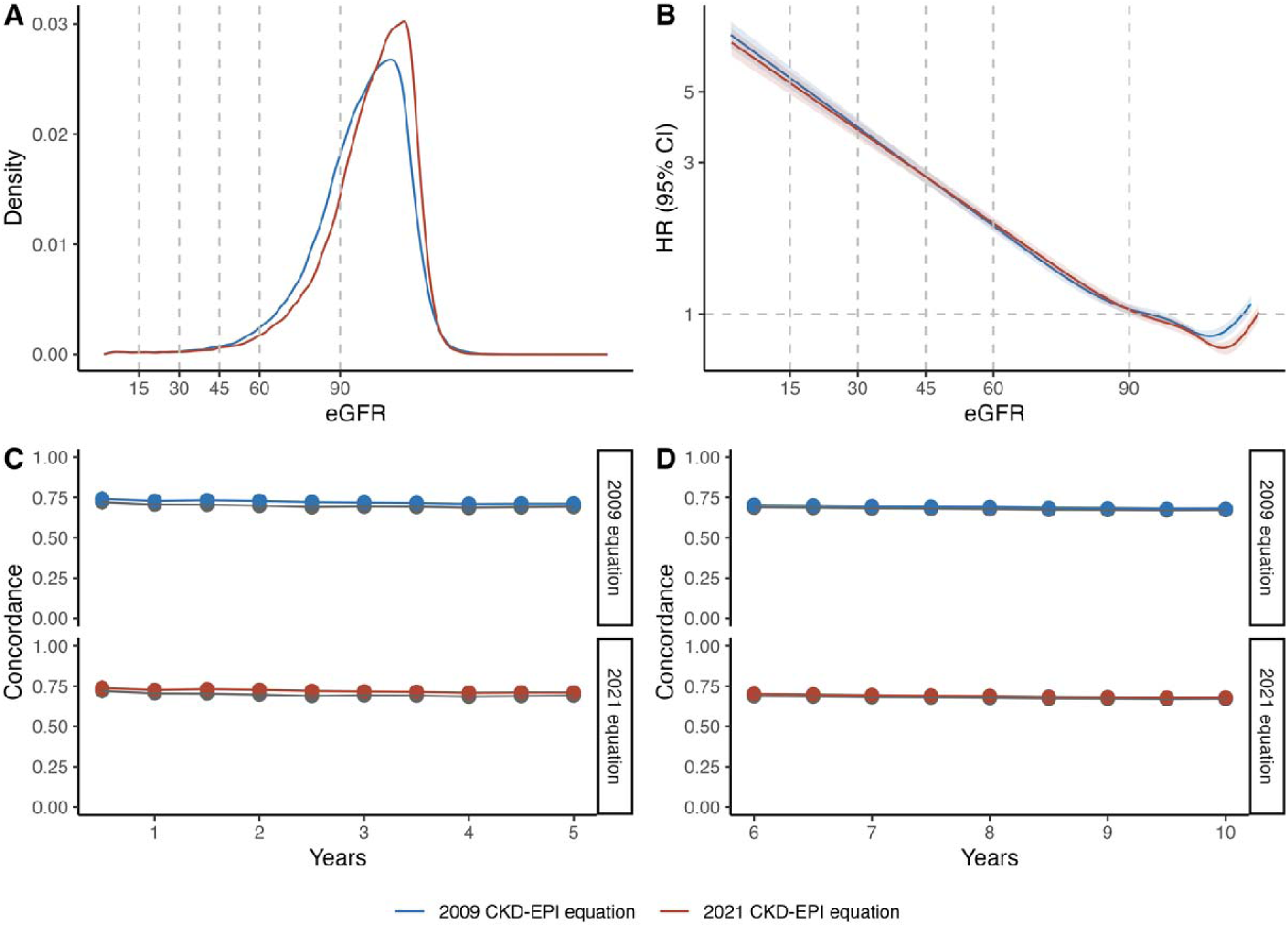
eGFR distribution and all-cause mortality risk prediction performance of eGFR estimated with either 2021 or 2009 CKD-EPI creatinine equation in the Mexico City Prospective Study. (A) Comparison of eGFR distribution in MCPS, (B) Comparison of HRs estimations across continuous eGFR levels, (C) 5-year discrimination assessment of eGFR estimated with either 2009 or 2021 CKD-EPI creatinine equation, (D) 10-year discrimination assessment of eGFR estimated with either 2009 or 2021 CKD-EPI creatinine equation. In both C and D, the gray line represents the baseline model without the eGFR covariate. All models were stratified by age category and adjusted for sex, place of residence, educational level, smoking, diabetes, history of cardiovascular disease, systolic blood pressure, and total cholesterol. To this baseline model, GFR estimated with either the 2021 or 2009 CKD-EPI creatinine equation was added. eGFR: Estimated glomerular filtration rate, CKD: Chronic Kidney Disease, CKD-EPI: Chronic Kidney Disease Epidemiology Collaboration.

**Table 2.** Discrimination assessment and overall performance assessment of all-cause, cardiovascular and kidney-related mortality for the baseline model and both extended models in participants of the Mexico City Prospective Study. The baseline model was stratified by age category and adjusted for sex, place of residence (Coyoacán or Iztapalapa), educational level (elementary, high school, university, other), smoking, diabetes, history of cardiovascular disease, systolic blood pressure, and total cholesterol. eGFR: Estimated glomerular filtration rate.

| Outcome | Model | Discrimination | 5-year overall performance |  | 10-year overall performance |  |
| --- | --- | --- | --- | --- | --- | --- |
|  |  | Harrell C-statistic (95%CI) | Brier score (95%CI) | Scaled Brier score | Brier score (95%CI) | Scaled Brier score |
| All-cause mortality | Baseline | 0.674 (0.669-0.68) | 0.036 (0.035–0.037) | 9.31% | 0.055 (0.054-0.056) | 26.62% |
|  | + eGFR 2009 | 0.690 (0.684-0.695) | 0.035 (0.034–0.036) | 12.77% | 0.053 (0.052-0.054) | 28.65% |
|  | + eGFR 2021 | 0.689 (0.684-0.695) | 0.035 (0.034–0.036) | 12.79% | 0.053 (0.052-0.054) | 28.66% |
| Cardiovascular mortality | Baseline | 0.672 (0.662-0.681) | 0.013 (0.013-0.014) | 5.41% | 0.027 (0.026-0.028) | 9.78% |
|  | + eGFR 2009 | 0.681 (0.671-0.690) | 0.013 (0.013-0.014) | 6.44% | 0.027 (0.026-0.028) | 10.46% |
|  | + eGFR 2021 | 0.680 (0.671-0.690) | 0.013 (0.013-0.014) | 6.44% | 0.027 (0.026-0.028) | 10.46% |
| Kidney-related mortality | Baseline | 0.834 (0.824-0.844) | 0.014 (0.013-0.014) | 1.61% | 0.029 (0.028-0.03) | 2.58% |
|  | + eGFR 2009 | 0.878 (0.869-0.887) | 0.014 (0.013-0.014) | 0.94% | 0.029 (0.029-0.03) | 1.83% |
|  | + eGFR 2021 | 0.878 (0.869-0.887) | 0.014 (0.013-0.014) | 0.92% | 0.029 (0.029-0.03) | 1.82% |

**Table 3.** Five- and ten-year risk of all-cause, cardiovascular, and kidney-related mortality, as well as age and sex-standardized outcome rates per 1,000 person-years related to reclassification of CKD-EPI 2009 eGFR categories with the CKD-EPI 2021 equation in participants of the Mexico City Prospective Study. All models were stratified by age category and adjusted for sex, place of residence, educational level (elementary, high school, university, other), smoking, diabetes, history of cardiovascular disease, systolic blood pressure, and total cholesterol. All-cause mortality was evaluated using Cox proportional hazard regression models, and cause-specific mortality using Fine & Gray sub distribution hazard models; reference group is No reclassification within the same category (G1-G2 and G3a-G5, respectively). *For all-cause mortality adjusted hazard ratios are presented, for cardiovascular and kidney-related mortality adjusted subdistribution hazard ratios are presented. eGFR: Estimated glomerular filtration rate. Abbreviations: HR, Hazard Ratio; CI, Confidence Interval; R, Reclassification; NR, No reclassification.

| Outcome | Reclassification with<br>CKD-EPI 2021 | 5-year |  |  | 10-year |  |  |
| --- | --- | --- | --- | --- | --- | --- | --- |
|  |  | Event rate<br>at 5 years<br>(R/NR) | HR* | 95% CI<sup>1</sup> | Event rate<br>at 10 years<br>(R/NR) | HR* | 95% CI<sup>1</sup> |
| All-cause mortality | G2 to G1 | 13.05 / 15.12 | 0.90 | 0.82, 0.97 | 15.89 / 17.47 | 0.93 | 0.88, 0.99 |
|  | Upward from G3a-G5 | 29.51 / 75.97 | 0.50 | 0.42, 0.59 | 29.76 / 63.43 | 0.60 | 0.53, 0.68 |
| Cardiovascular<br>mortality | G2 to G1 | 5.02 / 5.77 | 0.84 | 0.73, 0.96 | 5.82 / 6.46 | 0.93 | 0.85, 1.02 |
|  | Upward from G3a-G5 | 11.96 / 23.70 | 0.58 | 0.44, 0.76 | 12.46 / 18.19 | 0.69 | 0.56, 0.84 |
| Kidney-related<br>mortality | G2 to G1 | 0.86 / 0.95 | 0.88 | 0.63, 1.23 | 1.07 / 1.31 | 0.89 | 0.73, 1.10 |
|  | Upward from G3a-G5 | 6.54 / 32.06 | 0.33 | 0.22, 0.48 | 5.81 / 24.32 | 0.44 | 0.33, 0.58 |

### Prognostic significance of reclassification in eGFR categories

Using the 2021 equation, 11,838/142,884 participants (8.29%) were reclassified to higher eGFR categories compared to categories classified using the 2009 equation. Amongst them, 10,414/138,256 participants (7.53%) were reclassified from G2 to G1, and 1,061/4,628 (22.92%) were reclassified from G3a-G5 upward. Participants who were reclassified from categories G3a-G5 upward using the 2021 equation had a lower risk of all-cause, cardiovascular, and kidney-related mortality at 5 and 10 years compared to those not reclassified in the same categories. Similarly, participants classified upwards from G2 to G1 had a lower risk of 5- and 10-year all-cause, and 5-year cardiovascular mortality. No association was observed for cardiovascular mortality for those reclassified upward from G2 to G1 (**Figure 4**, **Table 3**).

**Figure 4.**
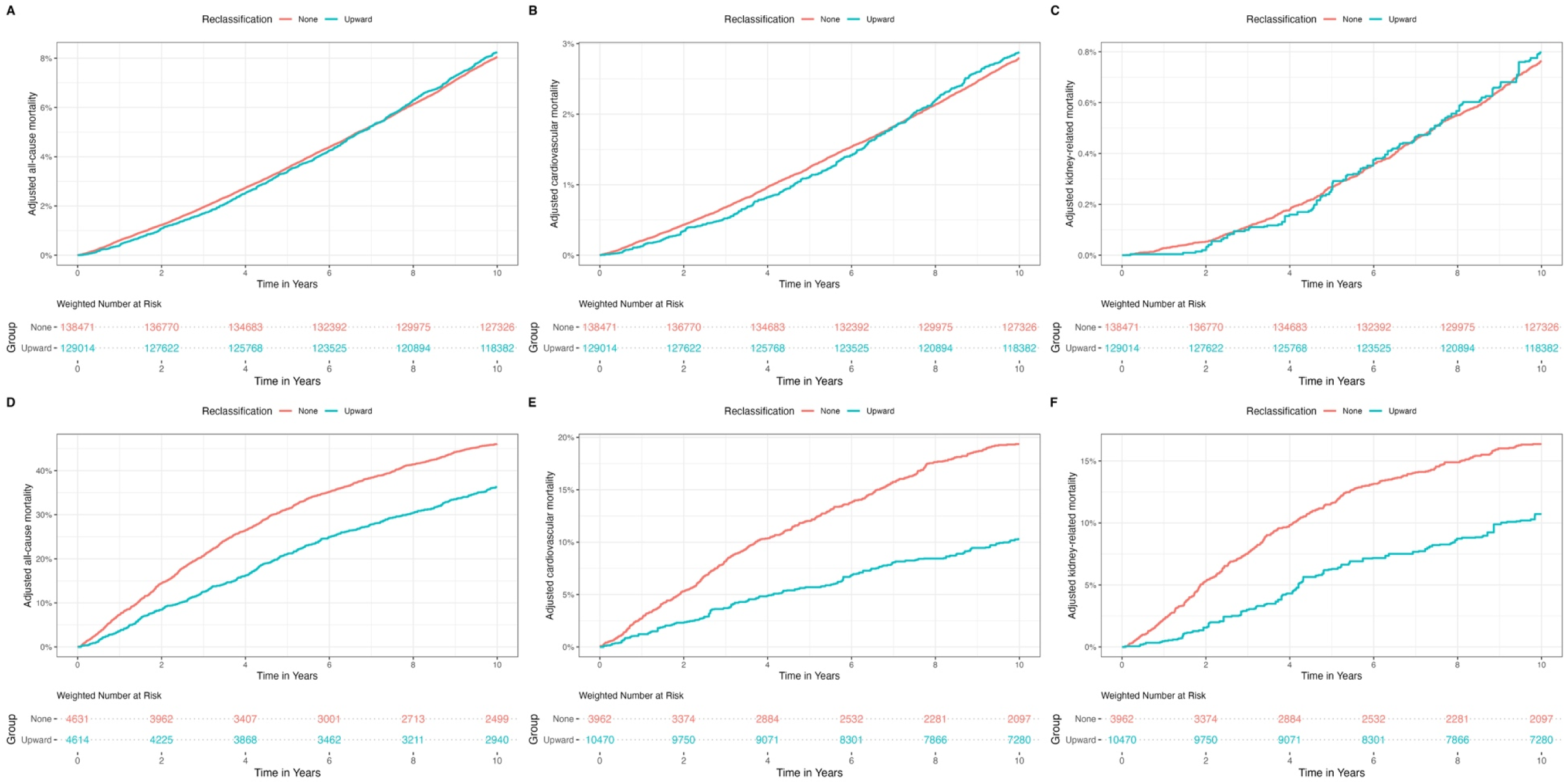
Kaplan-Meier cumulative incidence curves adjusted using inverse probability of treatment weighting (IPTW) by age, sex, community of origin, educational level, smoking, diabetes, prior cardiovascular disease, systolic blood pressure and total cholesterol for all-cause, cardiovascular and kidney-related mortality in MCPS participants. Panels A-C compare participants reclassified from G2 to G1 and Panels D-F participants reclassified from G3a-G5 upward using the 2021 eGFR creatinine-based CKD-EPI equation compared to non-reclassified participants in the same categories for risk of these outcomes.

## DISCUSSION

To our knowledge, this is the first study estimating the prevalence of eGFR categories and low eGFR using a nationally representative sample in Mexico. Our results using the 2021 CKD-EPI creatinine-based equation, show stable prevalence estimates of eGFR categories from 2016 to 2023. eGFR estimates were consistently lower using the 2009 CKD-EPI equation and, consequently, there was a pattern of upward reclassification adopting the newest 2021 CKD-EPI creatinine-based equation, particularly for older adults with comorbidities. Accordingly, CKD prevalence based only on low eGFR with the 2009 equation was higher compared to the 2021 equation. The predictive performance was similar for both equations; however, individuals reclassified to higher eGFR categories using the 2021 equation showed a lower risk of mortality outcomes compared to similar categories using the 2009 equation. Our results support the use of the 2021 equation to estimate GFR in Mexican population as it shows prognostically relevant reclassification of eGFR categories; however, we argue that the choice of equation should be made based on individual countries’ population structures, and on a wider array of kidney-related outcomes.

Previous reports have estimated CKD prevalence in Mexico relying on heterogeneous data sources and statistical methods^23,24^, or in specific geographical locations^25–28^. An analysis for the Global Burden of Disease estimated around 14.5 million prevalent cases of CKD in Mexico on 2017^1^, which contrast significantly with our estimations. Based solely on eGFR, we estimate that ∼2.25 million individuals are living with CKD in 2023, which increases to ∼2.4 million when considering prior CKD self-reported diagnosis in ENSANUT. These estimates of low eGFR prevalence in Mexico are lower compared to estimates for white adults in the US (2.9% vs. 6.3%), and similar to estimates from Hispanic adults (2.2%) derived from NHANES 2017-2020. This indicates either ethnic-specific differences in kidney function or CKD burden, or potential eGFR overestimation in adults of Hispanic descent. Further studies are warranted to provide more accurate estimates of CKD in Mexico, considering chronicity, albuminuria, cystatin-C measures, or the number of RRT users.

Recent clinical guidelines in the US have advocated for the implementation of eGFR equations without a race coefficient^2^. This recommendations has, however, raised concerns given that the 2021 CKD-EPI equation may lead to higher eGFR, and lower prevalence estimates in non-black individuals^6^. Since no specific equations have been developed for Hispanic of specifically for Mexican population, the use of eGFR equations without a race coefficient may lead to more precise estimates over prior alternatives. Similar to our estimations, previous studies in diverse populations have reported higher eGFR distribution, and lower CKD prevalence using the 2021 CKD-EPI equation, with upward reclassification especially among men, and older individuals^29–31^. Notably, Mexican clinical guidelines on CKD which were last updated in 2019 recommend GFR estimation with either Cockroft-Gault equation, the MDRD equation, or the 2009 CKD-EPI creatinine-based equation^32^. It should be noted that Cockroft-Gault is a creatinine clearance equation, and MDRD systematically underestimates higher GFR values^33^. We observed that the 2021 CKD-EPI equation yielded a similar but slightly improved prediction for all-cause, cardiovascular and kidney-related mortality compared to the 2009 equation, and reclassifies individuals at lower risk of these outcomes to higher eGFR categories. Our results support that the 2021 CKD-EPI creatinine-based equation should a reasonable choice for populations like Mexico, which is relatively homogeneous and comprised mainly of individuals with Hispanic descent. Additionally, creatinine measurements are widely used across the country to evaluate kidney function in clinical practice, and due to cost constraints, widespread implementation of cystatin C testing in Mexico remains a distant prospect.

Our study has several strengths. To our knowledge, this is the first study estimating GFR and CKD prevalence directly from creatinine measurements in Mexico, rather than relying on statistical methods to infer estimates. Additionally, given the data availability since 2016, we were able to obtain trends of eGFR and CKD estimates over the last six nationally representative surveys. Furthermore, access to data from a prospective study with mortality follow-up, allowed us to evaluate the predictive performance of eGFR equations for cause-specific outcomes in a predominately Hispanic population, broadening similar results in mostly white and European countries. However, some limitations should be considered to adequately interpret our results. First, CKD prevalence was estimated considering only low eGFR, and we were unable to include albuminuria to classify individuals as recommended by clinical guidelines^2^. Second, given the cross-sectional nature of the study, our definition of CKD relied on a single creatinine measurement, which lacks the three-month chronicity criterion, leading to a potential overestimation of CKD prevalence. Notably, our estimates did not account for the use of RRT. Moreover, due to the lack of data, we were unable to evaluate eGFR and CKD prevalence using the creatinine and cystatin C equation, which has been reported as the most accurate^6^. Finally, despite there being Afro-Mexican communities, its representation in ENSANUT and MCPS is uncertain. Given the admixture of Mexican population, most participants self-recognize as Mestizos^7^, and therefore neither ENSANUT nor MCPS routinely collect ethnicity data, which precluded us from being able to assess differences in prevalence and predictive performance for black Mexican adults.

In conclusion, eGFR and CKD prevalence based on low eGFR in Mexico have been stable over time, with some recent increases in 2023. As reported in predominately white Europeans, the new 2021 CKD-EPI equation consistently estimates higher GFR levels, which translates to lower CKD prevalence estimates and upward reclassification predominately in men and older individuals. Additionally, the 2021 equation yields a better predictive performance and adequately reclassified participants according to their risk for all-cause, cardiovascular and kidney-related morality compared to the 2009 equation. Therefore, we recommend implementing the creatinine-based 2021 CKD-EPI equation as a reliable choice for estimating eGFR and tracking CKD prevalence in Mexico.

## ACKNOWLEDGMENTS

This project was registered and approved by the Research Committee at Instituto Nacional de Geriatría, project number DI-PI-009-2024. JPE and CAFM are enrolled at the PECEM Program of the Faculty of Medicine at UNAM and are supported by CONACyT. JAS was supported by Grant Number K23DK135798 from the NIH/NIDDK and by the Massachusetts General Hospital Executive Committee and Center for Diversity and Inclusion Physician-Scientist Development Award. This research was conducted using Mexico City Prospective Study (MCPS) data obtained through an open-access data request (application number 2022-012). MCPS is a long-standing scientific collaboration between researchers at the National Autonomous University of Mexico and the University of Oxford, and has received funding from the Mexican Health Ministry, the National Council of Science and Technology for Mexico, Wellcome, Cancer Research UK, the British Heart Foundation, Kidney Research UK and the UK Medical Research Council. The authors thank the participants for their willingness to take part in this prospective study 20 years ago.

## AUTHOR CONTRIBUTIONS

Research idea and study design: DRG, CAFM, PSC, OYBC; data acquisition: CAFM, DRG, PSC, JAD, PKM, JBC, RTC, OYBC; analysis/interpretation: DRG, CAFM, OYBC; statistical analysis: DRG, CAFM, OYBC; manuscript drafting: DRG, CAFM, PSC, BGCF, JPE, JPDS, KBCH, LACQ, JCB, PKM, RTC, JAD, NEAV, JAS, OYBC; supervision or mentorship: OYBC. Each author contributed important intellectual content during manuscript drafting or revision and accepts accountability for the overall work by ensuring that questions pertaining to the accuracy or integrity of any portion of the work are appropriately investigated and resolved.

## DATA AVAILABILITY

All code and materials regarding ENSANUT are available for reproducibility of results http://github.com/oyaxbell/ckdepi_mcps_ensanut/. Data from the Mexico City Prospective Study are available to bona fide researchers. For more details, the study’s Data and Sample Sharing policy may be downloaded (in English or Spanish) from https://www.ctsu.ox.ac.uk/research/mcps. Available study data can be examined in detail through the study’s Data Showcase, available at https://datashare.ndph.ox.ac.uk/mexico/.

## ETHICAL DISCLOSURES

This project was registered and approved by the Research Committee at Instituto Nacional de Geriatría, project number DI-PI-009-2024. MCPS was approved by Ethics Committees at the Mexican Ministry of Health, the Mexican National Council for Science and Technology, and the University of Oxford, UK. All MCPS participants provided written informed consent.

