## Supplementary Material for "Comparison of low eGFR prevalence and prediction for mortality using 2009 and 2021 CKD-EPI equations in Mexican adults"

**SUPPLEMENTARY METHODS**

*Ethics approval in MCPS*

The study protocol of the Mexico City Prospective Study was approved by the corresponding ethics committees at the Mexican Ministry of Health, the Mexican National Council for Science and Technology, and the University of Oxford, and every study participant provided written informed consent.

*Creatinine measurements in MCPS*

Non-fasting blood samples were collected for all included participants for subsequent analysis upon evaluation by questionnaires and anthropometric measures. Samples were stored in a central laboratory, separated into plasma and buffy coat, and then frozen to –80ºC. From 2018 to 2024, every available plasma sample from the baseline examination was analyzed using nuclear magnetic resonance (NMR) spectroscopy at Nightingale Health Ltd (Kuopio, Finland) and the Clinical Trial Service Unit’s Wolfson Laboratory, quantifying over 249 plasma biomarkers including creatinine, as described previously^12^. Creatinine values obtained by NMR were converted to mg/dL using the formula (creatinine [mmol] x 1000) / 88.42.

*Mortality assessment in MCPS*

Mortality follow-up for participants was conducted through electronic probabilistic linkage to national death registries, with deaths tracked from the date of first assessment through September 30^th^, 2022. Deaths were coded according to the 10th Revision of the International Classification of Diseases (ICD-10). As mortality was determined via national registry linkage, follow-up is considered complete for all participants up to the date of death or last known follow-up. Participants who were alive at the end of the follow-up period were censored on September 30^th^, 2022. For the current analyses, all participants were censored at either five or ten years of follow-up to evaluate outcome risks and the predictive performance of various eGFR equations.

*Comorbidity assessment in MCPS and ENSANUT*

In ENSANUT, diabetes mellitus was defined as any participant with self-reported medical diagnosis of diabetes, or fasting plasma glucose of ≥126 mg/dL, or HbA1c ≥6.5%. Due to the lack of fasting plasma glucose measurement in MCPS, we defined diabetes as any participant with self-reported medical diagnosis of diabetes (including those reporting use of glucose-lowering medication), or HbA1c ≥6.5%^14^. In both datasets, hypertension was defined as any participant with either systolic (SBP) or diastolic blood pressure (DBP) ≥130/80 mmHg, and obesity as those with a calculated body mass index (BMI) ≥30 kg/m2.

*Complex survey design and weight calculation in ENSANUT*

ENSANUT uses probabilistic stratified multistage sampling for all its rounds. Stratified sampling for ENSANUT 2016 and 2018 considers per state, locality size, and sociodemographic characteristics, whilst ENSANUT 2020, 2021, 2022, and 2023 only consider state and locality size. Localities are categorized in most surveys according to the number of residents in metropolitan (capital cities or cities/metropolitan areas with ≥100,000 residents), urban/urban complement (areas with 2,500 – 999,999 residents not included in the metropolitan category), and rural (areas with <2,500 residents). For 2018, multiple sociodemographic strata were created based on 34 indicators which provide information on dwelling (e.g., overcrowding, access to basic services, etc.) and inhabitants (e.g., education level, employment, entitlement to health services, etc.) characteristics. The primary sampling units (PSUs) are AGEBs, which stand Basic Geostatistical Areas. They represent the fundamental unit of the National Geostatistical Framework, a system proposed by INEGI that divides Mexico’s territory into distinct levels of disaggregation. AGEBs are divided into urban and rural according to population density. Finally, sample selection is usually done per stages beginning with PSUs, localities, systematic sampling by pseudo-blocks, households and finally individuals. This complex survey design is considered by incorporating strata, PSUs, and sampling weights using the *survey* R package. The implemented sample weights were available for participants randomly subsampled for venous blood collection and were recalculated to account for missing data whenever this was observed in ENSANUT. Complex survey design was used to calculate weighted prevalence and 95% confidence intervals, as well as for estimating total event counts using the *survey* R package.

*Statistical Analyses*

***All-cause and cause-specific mortality analyses***

To assess performance of CKD-EPI equations for prediction of all-cause mortality we used Cox proportional hazard regression models stratified by age category at baseline and adjusted for sex, place of residence (Coyoacán or Iztapalapa), educational level (elementary, high school, university, other), smoking, diabetes, history of cardiovascular disease (previous self-reported diagnosis of angina, or myocardial infarction, or stroke), SBP, and total cholesterol (as continuous variables). For performance of cause-specific causes, including cardiovascular and kidney-related mortality, we fitted Fine & Gray semiparametric competing risk regression models using the *survival* R package. For restricted cubic spline analyses used in Cox and Fine & Gray models, knots for eGFR 2009 were located at 65.9, 89.8, 100.3, 106.9 and 119.5 mL/min/1.73m2, whilst for eGFR 2021 knots were located at 69.9, 94.19, 104.3, 111.9, and 120.6 mL/min/1.73m2. Non-linear splines were fitted using the *rms* R package. Outliers were not eliminated, unless they represented negative creatinine values, which were considered implausible and excluded from the analysis. Missing data analyses were conducted and found that missing data was more likely in older participants, residents of Iztapalapa, with higher educational level and higher burden of comorbidities (**Supplementary Table 6**); given these trends, we did not perform multiple imputation, as we consider that the MCAR assumption was not met. Therefore, complete case analyses were conducted in MCPS. In ENSANUT, missing data was minimal (**Supplementary Table 1**), and it was considered in the recalculation of sample weights. For all models, the proportional hazards assumption was tested using Schoenfeld residuals. Despite all failing to meet the proportional hazards assumption (p<0.001) largely due to extended follow-up and large sample sizes, upon visual inspection the proportional hazards assumption was largely met, as shown below in **Figures A-H.**

**Figure A.** Schoenfeld residuals for eGFR estimated with the 2009 equation smoothed using restricted cubic splines with five knots for prediction of all-cause 5-year mortality.

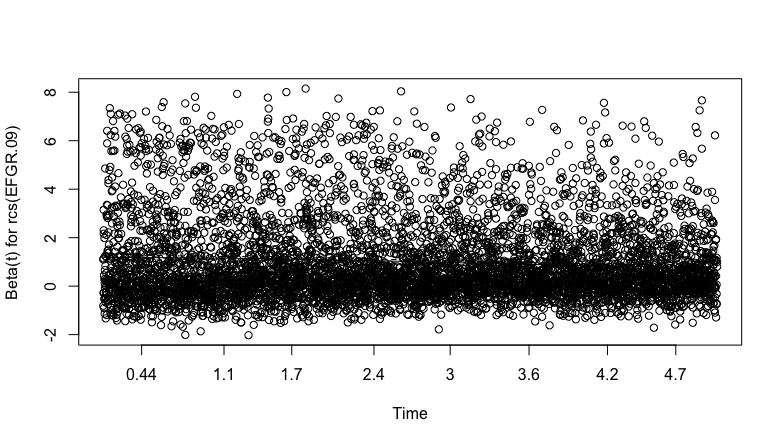

**Figure B.** Schoenfeld residuals for eGFR estimated with the 2021 equation smoothed using restricted cubic splines with five knots for prediction of all-cause 5-year mortality

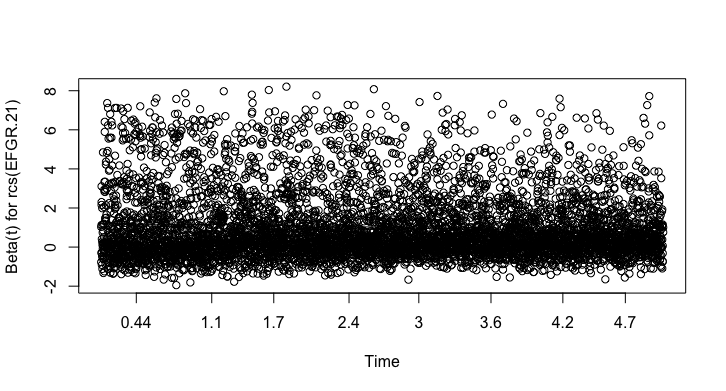

**Figure C.** Schoenfeld residuals for eGFR estimated with the 2009 equation smoothed using restricted cubic splines with five knots for prediction of all-cause 10-year mortality

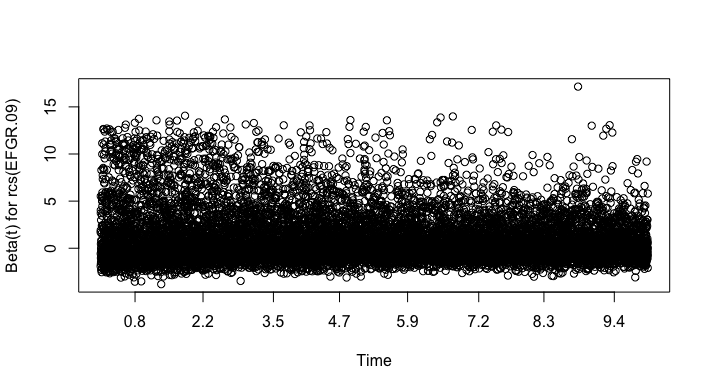

**Figure D.** Schoenfeld residuals for eGFR estimated with the 2021 equation smoothed using restricted cubic splines with five knots for prediction of all-cause 10-year mortality

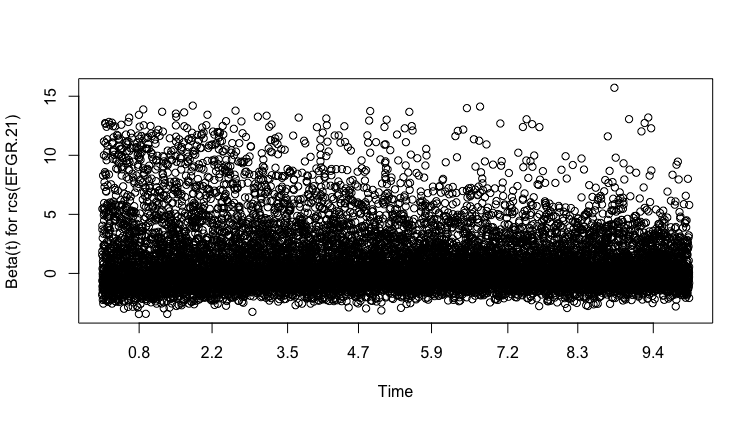

**Figure E.** Schoenfeld residuals for eGFR estimated with the 2009 equation smoothed using restricted cubic splines with five knots for prediction of CVD 5-year mortality

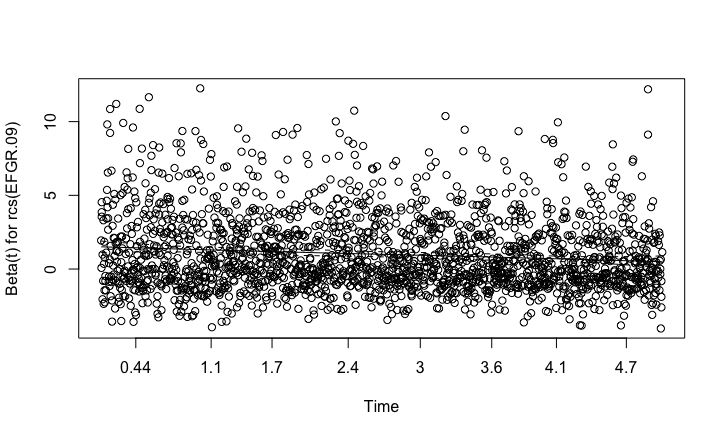

**Figure F.** Schoenfeld residuals for eGFR estimated with the 2021 equation smoothed using restricted cubic splines with five knots for prediction of CVD 5-year mortality

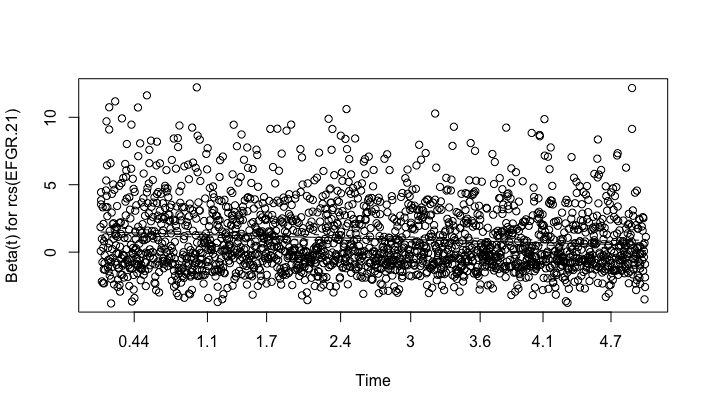

**Figure G.** Schoenfeld residuals for eGFR estimated with the 2009 equation smoothed using restricted cubic splines with five knots for prediction of CVD 10-year mortality

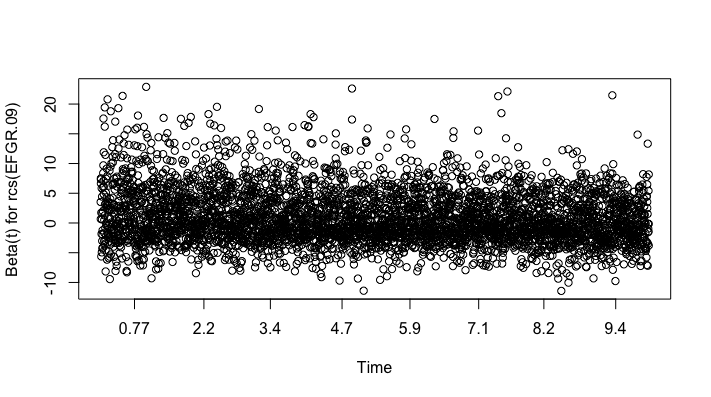

**Figure H.** Schoenfeld residuals for eGFR estimated with the 201 equation smoothed using restricted cubic splines with five knots for prediction of CVD 10-year mortality

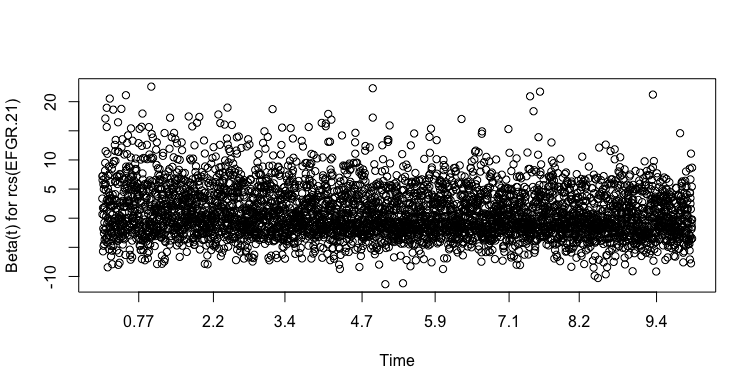

**SUPPLEMENTARY FIGURES**

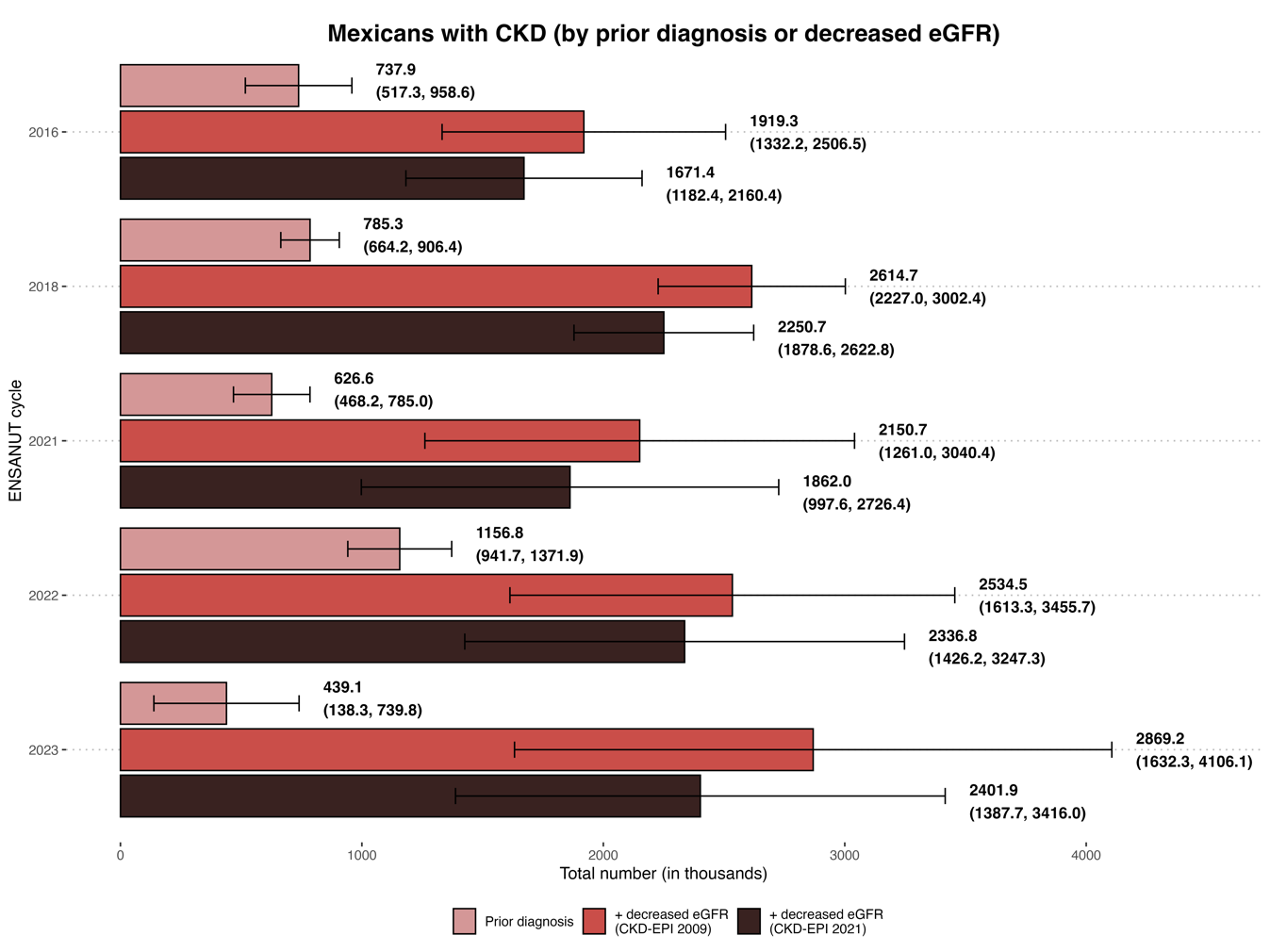

**Supplementary Figure 1.** Estimated total number of adults ≥20 years with self-reported CKD medical diagnosis, and the combination of previous CKD diagnosis or an eGFR threshold for CKD (eGFR <60 mL/min/1.73m2) using either CKD-EPI 2009 or 2021 creatinine equations in Mexico from 2016 to 2023 using ENSANUT data.

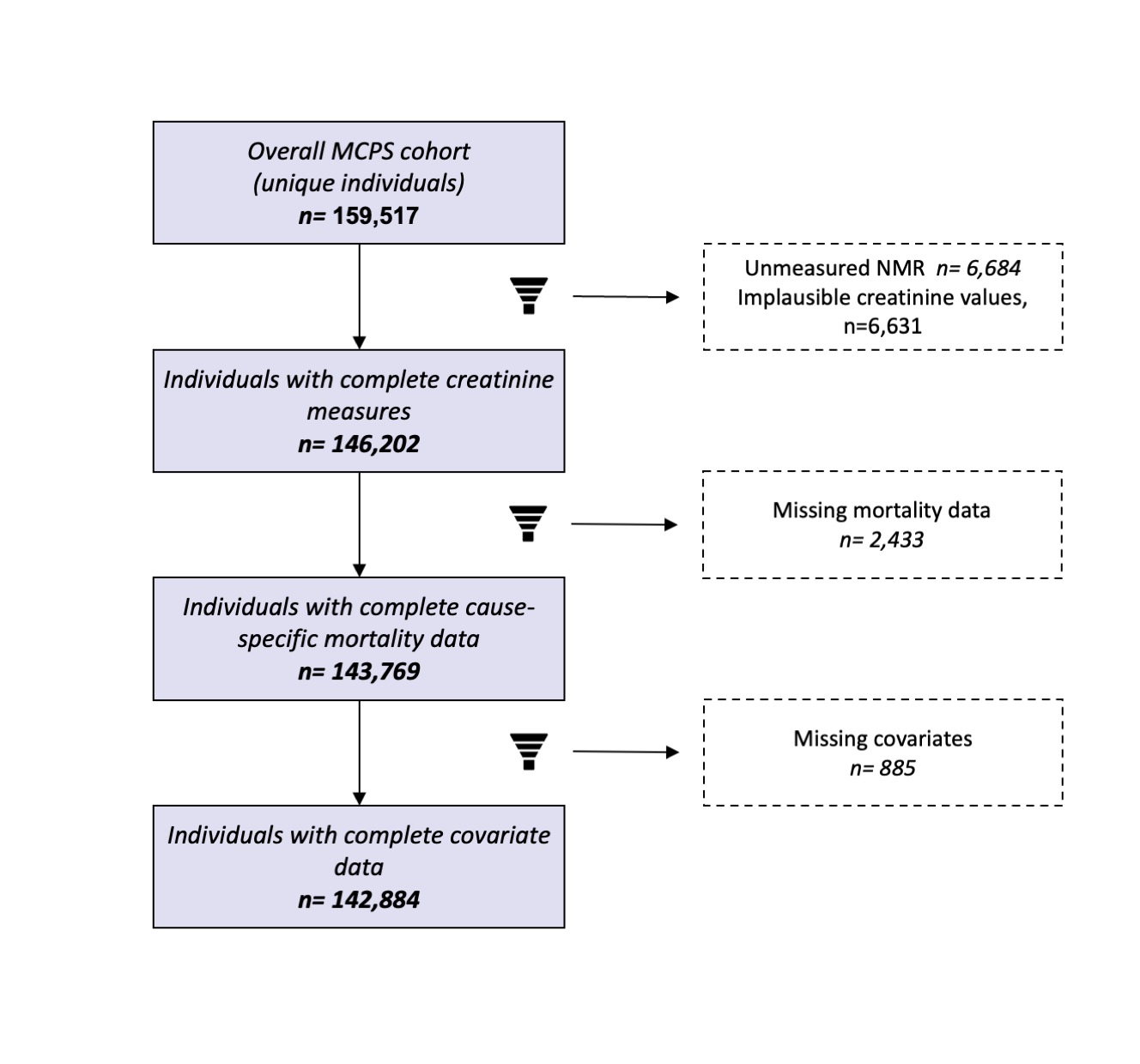

**Supplementary Figure 2.** Flowchart of participants from the Mexico City Prospective Study outlining reasons for exclusion from the present analysis

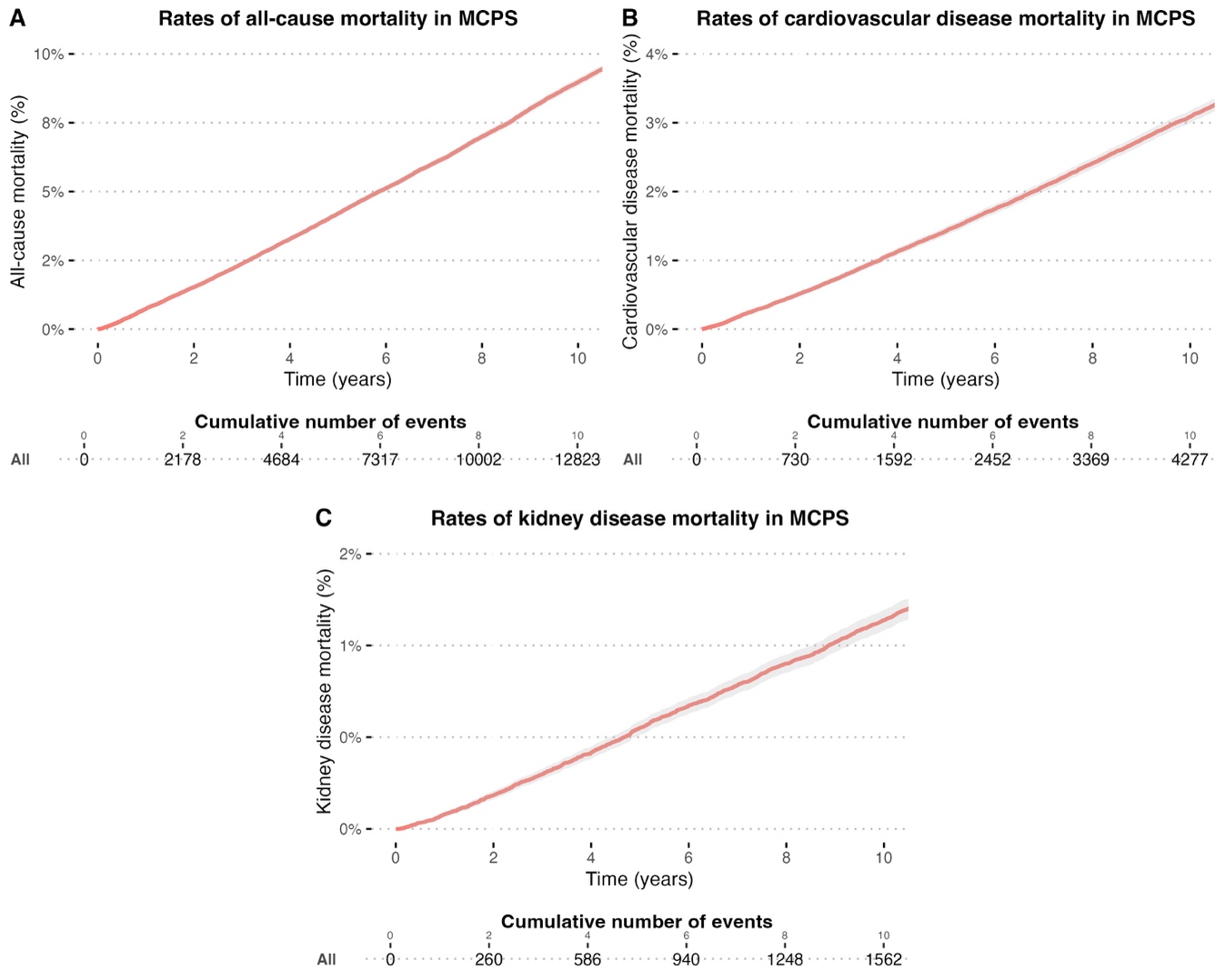

**Supplementary Figure 3.** Cumulative distribution curve showing rates of all-cause, cardiovascular, and kidney-related mortality over 10 years of follow-up in 142,884 participants ≥35 years of the Mexico City Prospective Study.

**SUPPLEMENTARY TABLES**

**Supplementary Table 1.** Characteristics of interviewed individuals according to ENSANUT survey cycle for the 2016-2023 period. Each survey shows the total number of participants in each cycle (n) along with the expanded population of individuals to whom these estimates are representative using ENSANUT complex survey design (N).

| **Characteristic** | **ENSANUT 2016** | **ENSANUT 2018** | **ENSANUT 2020** | **ENSANUT 2021** | **ENSANUT 2022** | **ENSANUT 2023** |
| --- | --- | --- | --- | --- | --- | --- |
|  | n = 3,930^1^  N= **70,383,884** | n = 13,155^1^  N= **82,710,347** | n = 2,347^1^  N= **83,869,561** | n = 2,152^1^  N= **84,829,009** | **n** = 2,091^1^  **N=** **85,520,767** | n = 1,435^1^  **N=** **76,477,260** |
| **Age (years)** | 44 (33, 58) | 44 (33, 58) | 45 (32, 58) | 45 (33, 59) | 47 (34, 60) | 47 (33, 60) |
| **Sex (women)** | 1,336 (34%) | 5,540 (42%) | 954 (41%) | 786 (37%) | 783 (37%) | 526 (37%) |
| *Missing* | 9 | 0 | 0 | 0 | 0 | 0 |
| **Smoking status** |  |  |  |  |  |  |
| Never | 2,079 (53%) | 8,310 (63%) | - | 1,441 (67%) | 1,398 (67%) | 950 (66%) |
| Former | 1,386 (35%) | 2,806 (21%) | - | 379 (18%) | 357 (17%) | 254 (18%) |
| Current | 460 (12%) | 2,008 (15%) | - | 326 (15%) | 332 (16%) | 226 (16%) |
| *Missing* | 15 | 31 | 2,346 | 6 | 4 | 131 |
| **Indigenous language** | 483 (12%) | 1,175 (8.9%) | 125 (5.3%) | 101 (4.7%) | 127 (6.1%) | 46 (3.2%) |
| **Urban area** | 1,779 (45%) | 8,542 (65%) | 1,794 (76%) | 1,595 (74%) | 1,531 (73%) | 1,027 (72%) |
| **Body mass index (kg/m^2)** | 28.0 (24.9, 31.7) | 28.4 (25.2, 31.9) | 28.3 (25.1, 32.2) | 28.8 (25.3, 32.7) | 28.9 (25.6, 32.4) | 28.9 (25.6, 33.2) |
| *Missing* | 103 | 338 | 138 | 47 | 49 | 32 |
| **Waist circumference (cm)** | 95 (87, 103) | 96 (87, 104) | - | 98 (89, 106) | 98 (89, 107) | 99 (90, 110) |
| *Missing* | 221 | 744 | 2,347 | 119 | 107 | 70 |
| **Fasting plasma glucose (mg/dL)** | 95 (88, 105) | 91 (83, 103) | 89 (82, 100) | 93 (86, 104) | 90 (81, 102) | 93 (85, 105) |
| *Missing* | 0 | 1 | 0 | 0 | 0 | 0 |
| **HbA1c (%)** | 5.40 (5.10, 5.70) | 5.30 (5.00, 5.70) | 5.40 (5.10, 5.80) | 5.50 (5.20, 5.90) | 5.40 (5.10, 5.90) | 5.40 (5.10, 5.90) |
| *Missing* | 140 | 298 | 15 | 23 | 37 | 21 |
| **Total cholesterol (mg/dL)** | 185 (161, 211) | 182 (159, 209) | 176 (152, 203) | 176 (152, 202) | 151 (122, 182) | 148 (123, 176) |
| **Triglycerides (mg/dL)** | 165 (114, 241) | 167 (115, 243) | 152 (106, 223) | 149 (107, 217) | 131 (91, 192) | 126 (90, 186) |
| **Creatinine (mg/L)** | 0.68 (0.58, 0.81) | 0.68 (0.57, 0.82) | 0.69 (0.58, 0.80) | 0.70 (0.60, 0.80) | 0.66 (0.54, 0.80) | 0.69 (0.59, 0.84) |
| **Hypertension (%)** | 1,029 (27%) | 4,342 (37%) | 692 (31%) | 639 (30%) | 700 (34%) | 415 (40%) |
| *Missing* | 96 | 1,295 | 101 | 33 | 25 | 401 |
| **Type 2 diabetes (%)** | 658 (17%) | 2,394 (19%) | 450 (19%) | 420 (20%) | 448 (22%) | 306 (22%) |
| *Missing* | 118 | 249 | 13 | 18 | 29 | 18 |
| **Cardiovascular disease (%)** | 123 (3.1%) | 383 (2.9%) | 45 (1.9%) | 36 (1.7%) | 88 (4.2%) | 55 (3.8%) |
| *Missing* | 5 | 0 | 0 | 0 | 0 | 0 |
| **Prior CKD (%)** | 48 (1.2%) | 140 (1.1%) | - | 9 (0.4%) | 29 (1.4%) | 18 (1.3%) |
| *Missing* | 5 | 0 | 2,347 | 0 | 0 | 0 |
| **eGFR 2009 (mL/min/1.73m2)** | 108 (94, 119) | 109 (95, 120) | 109 (96, 121) | 107 (94, 119) | 109 (96, 122) | 106 (92, 118) |
| *Missing* | 9 | 0 | 0 | 0 | 1 | 0 |
| **eGFR 2021 (mL/min/1.73m2)** | 111 (99, 121) | 111 (99, 122) | 112 (100, 123) | 110 (98, 121) | 111 (100, 122) | 109 (96, 120) |
| *Missing* | 9 | 0 | 0 | 0 | 1 | 0 |
| ^1^Median (IQR); n (%) | | | | | | |

| **Year** | **2021 CKD-EPI equation** | **2009 CKD-EPI equation** | **Difference** |
| --- | --- | --- | --- |
| **2016** | 1,264,499.79 (822,068.21-1,706,931.38) | 1,512,462.04 (977,504.70-2,047,419.38) | -247,962.3 |
| **2018** | 1,630,610.30 (1,316,170.65-1,945,049.96) | 2,000,976.34 (1,666,825.65-2,335,127.03) | -370,366.0 |
| **2020** | 1,563,809.42 (1,065,219.95-2,062,398.89) | 1,975,743.18 (1,407,743.61-2,543,742.75) | -411,933.8 |
| **2021** | 1,568,795.96 (773,718.08-2,363,873.85) | 1,857,433.60 (1,032,624.60-2,682,242.60) | -288,637.6 |
| **2022** | 1,568,860.17 (820,977.95-2,316,742.40) | 1,766,582.00 (1,004,540.76-2,528,623.25) | -197,721.8 |
| **2023** | 2,251,124.88 (1,240,552.83-3,261,696.94) | 2,737,657.37 (1,501,064.90-3,974,249.83) | -486,532.5 |

**Supplementary Table 2.** Total number of individuals estimated to have CKD (eGFR <60mL/min/1.73m2) in Mexico from 2016 to 2023 using ENSANUT data with either the 2021 or 2009 CKD-EPI equation to estimate eGFR. eGFR: Estimated glomerular filtration rate, CKD-EPI: Chronic Kidney Disease Epidemiology Collaboration.

| **eGFR categories** | | | | | | |
| --- | --- | --- | --- | --- | --- | --- |
| **eGFR category** | **2016** | **2018** | **2020** | **2021** | **2022** | **2023** |
| **G1** | 84.72%  (82.70-86.74) | 82.92%  (81.70-84.14) | 85.24%  (83.49-86.99) | 82.14%  (79.67-84.60) | 84.67%  (82.23-87.10) | 78.64%  (75.94-81.34) |
| **G2** | 13.13%  (11.19-15.06) | 14.66%  (13.50-15.81) | 12.38%  (10.78-13.98) | 15.35%  (13.00-17.71) | 13.25%  (10.86-15.63) | 17.60%  (15.13-20.08) |
| **G3a** | 1.13%  (0.65-1.61) | 1.30%  (1.04-1.56) | 1.43%  (0.90-1.95) | 1.42%  (0.71-2.12) | 0.95%  (0.53-1.37) | 1.92%  (1.05-2.78) |
| **G3b** | 0.53%  (0.26-0.79) | 0.69%  (0.43-0.95) | 0.43%  (0.09-0.77) | 0.25%  (0.05-0.45) | 0.40%  (0.12-0.68) | 1.01%  (0.37-1.66) |
| **G4** | 0.14%  (0.02-0.27) | 0.28%  (0.16-0.41) | 0.30%  (0.09-0.52) | 0.32%  (0.00-0.67) | 0.55%  (0.00-1.26) | 0.24%  (0.00-0.59) |
| **G5** | 0.35%  (0.00-0.71) | 0.15%  (0.07-0.23) | 0.22%  (0.05-0.39) | 0.53%  (0.00-1.33) | 0.18%  (0.01-0.36) | 0.59%  (0.14-1.05) |
| **Low eGFR** | | | | | | |
| **eGFR <60 mL/min 1.73 m^2^** | 2.2%  (1.44–2.96) | 2.4%  (2.00–2.80) | 2.4%  (1.71–3.09) | 2.5%  (1.39–3.61) | 2.1%  (1.21–2.99) | 3.6  (1.99–5.21) |

**Supplementary Table 3.** Nationally representative eGFR categories prevalence from 2016 to 2023 using ENSANUT data. **eGFR was estimated with CKD-EPI 2009 creatinine-based equation**. G1 normal or high (≥90 mL/min per 1.73 m^2^), G2 mildly decreased (60–89 mL/min/1.73 m^2^), G3a mildly to moderately decreased (45–59 mL/min/1.73 m^2^), G3b moderately to severely decreased (30–44 mL/min/1.73 m^2^), G4 severely decreased (15-29 mL/min/1.73 m^2^), and G5 kidney failure (<15 mL/min/1.73 m^2^). eGFR: Estimated glomerular filtration rate, CKD-EPI: Chronic Kidney Disease Epidemiology Collaboration.

| **eGFR categories** | | | | | | |
| --- | --- | --- | --- | --- | --- | --- |
| **eGFR category** | **2016** | **2018** | **2020** | **2021** | **2022** | **2023** |
| **G1** | 82.59%  (80.07-85.12) | 81.44%  (79.98-82.91) | 84.84%  (82.73-86.95) | 80.04%  (77.10-82.99) | 84.30%  (81.71-86.89) | 81.62%  (77.88-85.36) |
| **G2** | 14.51%  (12.18-16.85) | 15.58%  (14.21-16.95) | 12.27%  (10.35-14.18) | 17.59%  (14.77-20.41) | 12.90%  (10.49-15.32) | 14.04%  (11.06-17.01) |
| **G3a** | 1.54%  (0.88-2.21) | 1.60%  (1.22-1.98) | 1.50%  (0.84-2.17) | 1.64%  (0.73-2.55) | 1.11%  (0.56-1.66) | 2.60%  (1.14-4.06) |
| **G3b** | 0.61%  (0.23-0.98) | 0.79%  (0.45-1.12) | 0.70%  (0.17-1.23) | 0.46%  (0.02-0.90) | 0.61%  (0.19-1.04) | 1.32%  (0.33-2.32) |
| **G4** | 0.23%  (0.02-0.44) | 0.39%  (0.21-0.57) | 0.34%  (0.05-0.64) | 0.27%  (0.00-0.69) | 0.80%  (0.00-1.88) | 0.01%  (0.00-0.02) |
| **G5** | 0.51%  (0.00-1.10) | 0.21%  (0.09-0.33) | 0.34%  (0.08-0.61) | 0.00%  (0.00-0.00) | 0.28%  (0.02-0.54) | 0.41%  (0.00-0.88) |

**Supplementary Table 4.** Nationally representative eGFR categories prevalence from 2016 to 2023 for adults ≥35 years using ENSANUT data. **eGFR was estimated with CKD-EPI 2021 creatinine-based equation**. G1 normal or high (≥90 mL/min per 1.73 m^2^), G2 mildly decreased (60–89 mL/min/1.73 m^2^), G3a mildly to moderately decreased (45–59 mL/min/1.73 m^2^), G3b moderately to severely decreased (30–44 mL/min/1.73 m^2^), G4 severely decreased (15-29 mL/min/1.73 m^2^), and G5 kidney failure (<15 mL/min/1.73 m^2^). eGFR: Estimated glomerular filtration rate, CKD-EPI: Chronic Kidney Disease Epidemiology Collaboration.

| **CKD proxy Prevalence** | | | | | |
| --- | --- | --- | --- | --- | --- |
|  | **2016** | **2018** | **2021** | **2022** | **2023** |
| **Prior CKD diagnosis** | 1.06%  (0.75-1.38) | 0.95%  (0.80-1.10) | 0.74%  (0.56-0.92) | 1.35%  (1.10-1.60) | 0.57%  (0.17-0.98) |
| **Prior DX + CKD-EPI 2009** | 2.73%  (1.89-3.58) | 3.16%  (2.70-3.63) | 2.91%  (1.71-4.10) | 2.99%  (1.92-4.07) | 3.75%  (2.13-5.37) |
| **Prior DX + CKD-EPI 2021** | 2.38%  (1.67-3.09) | 2.72%  (2.27-3.17) | 2.52%  (1.36-3.67) | 2.76%  (1.70-3.82) | 3.14%  (1.79-4.49) |
| **CKD total** | | | | | |
|  | **2016** | **2018** | **2021** | **2022** | **2023** |
| **Prior CKD diagnosis** | 737,916.60 (517,264.43-958,568.77) | 785,285.00 (664,169.24-906,400.76) | 626,590.40 (468,180.33-785,000.47) | 1,156,805.90 (941,672.37-1,371,939.43) | 439,092.20 (138,344.40-739,840.00) |
| **Prior DX + CKD-EPI 2009** | 1,919,342.60 (1,332,207.49-2,506,477.71) | 2614740.70 (2227047.07-3002434.33) | 2150671.00 (1260975.22-3040366.78) | 2534490.90 (1613254.32-3455727.48) | 2,869,199.20 (1,632,256.58-4,106,141.82) |
| **Prior DX + CKD-EPI 2021** | 1,671,380.30 (1,182,367.65-2,160,392.95) | 2,250,666.50 (1,878,560.67-2,622,772.33) | 1,862,033.40 (997,617.01-2,726,449.79) | 2,336,769.10 (1,426,194.41-3,247,343.79) | 2,401,866.70 (1,387,738.07-3,415,995.33) |

**Supplementary Table 5.** Prevalence estimations and total number of individuals with self-reported CKD medical diagnosis, and the combination of previous CKD diagnosis or low eGFR (<60 mL/min/1.73m^2^) with either CKD-EPI 2009 or 2021 creatinine-based equations in Mexico from 2016 to 2023, using ENSANUT data.

| **Characteristic** | **No reclassification**  N = 71,648,593 | **Upward reclassification**  N = 4,856,049 |
| --- | --- | --- |
| **Mean age** | 43 (42, 45) | 64 (57, 70) |
| **Age category** | | |
| <60 years | 85% (82%, 88%) | 31% (16%, 50%) |
| ≥60 years | 15% (12%, 18%) | 69% (50%, 84%) |
| **Creatinine** | 0.76 (0.72, 0.80) | 0.91 (0.83, 1.0) |
| **eGFR 2009** | 107 (105, 109) | 82 (78, 86) |
| **eGFR 2021** | 109 (108, 111) | 86 (82, 90) |
| **Sex** | | |
| Women | 54% (48%, 60%) | 32% (20%, 46%) |
| Men | 46% (40%, 52%) | 68% (54%, 80%) |
| **Diabetes** | | |
| No diabetes | 83% (79%, 86%) | 64% (49%, 76%) |
| Diabetes | 17% (14%, 21%) | 36% (24%, 51%) |
| **Hypertension** | | |
| No hypertension | 66% (59%, 71%) | 33% (18%, 52%) |
| Hypertension | 34% (29%, 41%) | 67% (48%, 82%) |
| eGFR: Estimated Glomerular Filtration Rate. | | |

**Supplementary Table 6.** Characteristics of reclassified individuals and those who were not reclassified using the 2021 CKD-EPI equation across every eGFR category using ENSANUT 2023 data. eGFR: Estimated glomerular filtration rate, CKD-EPI: Chronic Kidney Disease Epidemiology Collaboration.

| **Characteristic** | | **Overall**  N = 159,517*^1^* | **Non-missing data**  N = 142,884*^1^* | **Missing data**  N = 16,633*^1^* | **p-value***^2^* |
| --- | --- | --- | --- | --- | --- |
| **eGFR 2009 (mL/min/1.73m2)** | | 100 (88, 110) | 100 (88, 110) | 95 (83, 105) | <0.001 |
| *Missing* | | 13,315 | 0 | **13,315** |  |
| **eGFR 2021 (mL/min/1.73m2)** | | 104 (93, 113) | 104 (93, 113) | 99 (88, 108) | <0.001 |
| *Missing* | | 13,315 | 0 | **13,315** |  |
| **Age (years)** | | 53 (±13) | 52 (±13) | 55 (±14) | <0.001 |
| **Sex (%)** | |  |  |  | 0.6 |
| Female | | 106,938 (67%) | 95,820 (67%) | 11,118 (67%) |  |
| Male | | 52,579 (33%) | 47,064 (33%) | 5,515 (33%) |  |
| **Place of residence (%)** | |  |  |  | <0.001 |
| Iztapalapa | | 95,767 (60%) | 85,138 (60%) | 10,629 (64%) |  |
| Coyoacan | | 63,750 (40%) | 57,746 (40%) | 6,004 (36%) |  |
| **Education level (%)** | |  |  |  | <0.001 |
| University | | 24,399 (15%) | 21,446 (15%) | 2,953 (18%) |  |
| High school | | 38,529 (24%) | 34,849 (24%) | 3,680 (22%) |  |
| Elementary | | 74,918 (47%) | 67,489 (47%) | 7,429 (45%) |  |
| Other | | 21,587 (14%) | 19,100 (13%) | 2,487 (15%) |  |
| *Missing* | | 84 | 0 | 84 |  |
| **All-cause mortality (%)** | | 31,397 (20%) | 28,475 (20%) | 2,922 (21%) | 0.006 |
| *Missing* | | 2,649 | 0 | 2,649 |  |
| **Cardiovascular mortality (%)** | | 10,456 (6.7%) | 9,472 (6.6%) | 984 (7.0%) | 0.065 |
| *Missing* | | 2,649 | 0 | 2,649 |  |
| **Kidney disease mortality (%)** | | 3,344 (2.1%) | 3,060 (2.1%) | 284 (2.0%) | 0.4 |
| *Missing* | | 2,649 | 0 | 2,649 |  |
| **Diabetes(%)** | | 29,943 (19%) | 26,677 (19%) | 3,266 (20%) | 0.003 |
| **Hypertension** | | 34,002 (21%) | 30,174 (21%) | 3,828 (23%) | <0.001 |
| **CKD (%)** | | 1,364 (0.9%) | 1,209 (0.8%) | 155 (0.9%) | 0.3 |
| **Cardiovascular disease (%)** | | 4,281 (2.7%) | 3,717 (2.6%) | 564 (3.4%) | <0.001 |
| **HbA1c (%)** | | 5.54 (5.26, 5.99) | 5.54 (5.26, 5.99) | 5.63 (5.35, 6.18) | <0.001 |
| *Missing* | | 4,820 | 0 | 4,820 |  |
|  | *^1^*Median (Q1, Q3); Mean (±SD); n (%) | | | | |
|  | *^2^*Wilcoxon rank sum test; Pearson's Chi-squared test | | | | |

**Supplementary Table 7.** Comparison of cases with and without missing data in the MCPS dataset. **Abbreviations:** eGFR: Estimated glomerular filtration rate, CKD: Chronic Kidney Disease, CKD-EPI: Chronic Kidney Disease Epidemiology Collaboration, MCPS: Mexico City Prospective Study.

**Mexico City Prospective Study (MCPS) Supplementary Results**

eGFR and CKD prevalence

Using creatinine values obtained from baseline blood samples in the MCPS dataset, eGFR distribution was higher using the 2021 CKD-EPI equation compared to the 2009 CKD-EPI equation (**Figure 3A**). Correspondingly, and similar to our findings in ENSANUT, eGFR categories and CKD prevalence were lower using the CKD-EPI 2021 equation (**Supplementary Table 8**), with a CKD prevalence of 2.50% using the CKD-EPI 2021 equation and 3.24% using the CKD-EPI 2009 equation. Likewise, 0.85% of participants had a self-reported previous diagnosis of CKD, and defining CKD as the combination of previous self-reported medical diagnosis of CKD or decreased eGFR, 3.88% had CKD using the CKD-EPI 2009 equation, and 3.15% using the CKD-EPI 2021 (**Supplementary Table 9**).

| **eGFR category** | **CKD-EPI 2009 equation** | **CKD-EPI 2021 equation** |
| --- | --- | --- |
| **G1** | 103177 (72.21%) | 113591 (79.50%) |
| **G2** | 35079 (24.55%) | 25726 (18.00%) |
| **G3a** | 2899 (2.03%) | 2112 (1.48%) |
| **G3b** | 949 (0.66%) | 739 (0.52%) |
| **G5** | 438 (0.31%) | 399 (0.28%) |
| **G4** | 342 (0.24%) | 317 (0.22%) |
| **CKD** | 4628 (3.24%) | 3567 (2.50%) |

**Supplementary Table 8.** eGFR categories and CKD prevalence in the MCPS dataset using baseline blood samples estimated with both CKD-EPI equations. eGFR: Estimated glomerular filtration rate, CKD: Chronic Kidney Disease, CKD-EPI: Chronic Kidney Disease Epidemiology Collaboration, MCPS: Mexico City Prospective Study.

| **Previous diagnosis** | 1209 (0.85%) |
| --- | --- |
| **Previous diagnosis + eGFR 2009** | 5548 (3.88%) |
| **Previous diagnosis + eGFR 2021** | 4501 (3.15%) |

**Supplementary Table 9.** Prevalence estimations in MCPS dataset of individuals with self-reported CKD medical diagnosis, and the combination of previous CKD diagnosis or decreased eGFR using either CKD-EPI 2009 or CKD-EPI 2021 creatinine equations in Mexico from 2016 to 2023 using ENSANUT data. eGFR: Estimated glomerular filtration rate, CKD: Chronic Kidney Disease, CKD-EPI: Chronic Kidney Disease Epidemiology Collaboration, MCPS: Mexico City Prospective Study.

| **Outcome** | **Time-range** | **Model** | **Chi-squared statistic** | **LR test vs. baseline** | **Log-likelihood** | **p-value and LR test**  **2009 vs. 2021** |
| --- | --- | --- | --- | --- | --- | --- |
| **All-cause mortality** | **5-year** | **Baseline** | 1957.53 | - | -57603 | - |
|  |  | **+ eGFR 2009** | 3599.43 | 1642.9 | -56782 | LR test= 1.41  p<0.001  **Favours 2021** |
|  |  | **+ eGFR 2021** | **3600.84** | 1643.3 | -55782 |  |
|  | **10-year** | **Baseline** | 3632.27 | - | -123145 | - |
|  |  | **+ eGFR 2009** | **5194.90** | 1562.6 | -122363 | LR test= 0.61  P<0.001  **Favours 2009** |
|  |  | **+ eGFR 2021** | 5194.29 | 1562.0 | -122364 |  |
| **Cardiovascular mortality** | **5-year** | **Baseline** | 722.82 | - | -19019 | - |
|  |  | **+ eGFR 2009** | 1069.50 | 346.67 | -18846 | LR test= 2.77  p<0.001  **Favours 2021** |
|  |  | **+ eGFR 2021** | **1072.27** | 349.44 | -18844 |  |
|  | **10-year** | **Baseline** | 1256.40 | - | -40557 | - |
|  |  | **+ eGFR 2009** | 1585.94 | 329.54 | -40392 | LR test= 3.65  p<0.001  **Favours 2021** |
|  |  | **+ eGFR 2021** | **1589.60** | 333.2 | -40392 |  |
| **Kidney-related mortality** | **5-year** | **Baseline** | 1222.10 | - | -7057.7 | - |
|  |  | **+ eGFR 2009** | 2785.45 | 1563.4 | -6276 | LR test= 2.18  p<0.001  **Favours 2021** |
|  |  | **+ eGFR 2021** | **2787.63** | 1565.5 | -6274.9 |  |
|  | **10-year** | **Baseline** | 2337.42 | - | -14343 | - |
|  |  | **+ eGFR 2009** | **3972.61** | 1635.2 | -13526 | LR test= 1.070  p<0.001  **Favours 2009** |
|  |  | **+ eGFR 2021** | 3971.54 | 1643.1 | -13526 |  |

**Supplementary Table 10.** Likelihood ratio, log-likelihood and nested testing of models for of all-cause, cardiovascular and kidney-related mortality for the baseline model and both extended models in participants of the Mexico City Prospective Study. The baseline model was stratified by age category and adjusted for sex, place of residence (Coyoacán or Iztapalapa), educational level (elementary, high school, university, other), smoking, diabetes, history of cardiovascular disease, systolic blood pressure, and total cholesterol. eGFR: Estimated glomerular filtration rate.

| **Characteristic** | **HR**^1^ | **95% CI**^1^ | **p-value** |
| --- | --- | --- | --- |
| **Male sex** | 1.57 | 1.47, 1.66 | <0.001 |
| **Residence in Coyoacán** | 0.74 | 0.70, 0.78 | <0.001 |
| **Educational level** |  |  |  |
| University | Ref. | Ref. |  |
| High school | 1.16 | 1.02, 1.32 | 0.025 |
| Elementary | 1.38 | 1.24, 1.55 | <0.001 |
| Other | 1.46 | 1.29, 1.65 | <0.001 |
| **Smoking** |  |  |  |
| Never | Ref. | Ref. |  |
| Former | 1.14 | 1.07, 1.22 | <0.001 |
| Current | 1.15 | 1.07, 1.23 | <0.001 |
| **Diabetes** | 2.07 | 1.97, 2.19 | <0.001 |
| **Cardiovascular disease** | 1.67 | 1.53, 1.83 | <0.001 |
| **Systolic blood pressure (per mmHg)** | 1.00 | 1.00, 1.00 | <0.001 |
| **Total cholesterol (per mmol/L)** | 0.88 | 0.85, 0.90 | <0.001 |
| **eGFR estimated using the 2009 equation** |  |  |  |
| rcs(EFGR.09) | 0.97 | 0.97, 0.97 | <0.001 |
| rcs(EFGR.09)' | 1.04 | 1.03, 1.05 | <0.001 |
| rcs(EFGR.09)'' | 0.73 | 0.62, 0.85 | <0.001 |
| rcs(EFGR.09) ''' | 2.95 | 1.87, 4.64 | <0.001 |
| ^1^HR = Hazard Ratio, CI = Confidence Interval | | | |

**Supplementary Table 11.** Cox proportional hazard model for prediction of **all-cause mortality** at **5-years** using MCPS data and the **2009 CKD-EPI equation** for eGFR. The baseline model was stratified by age category and adjusted for sex, place of residence (Coyoacán or Iztapalapa), educational level (elementary, high school, university, other), smoking, diabetes, history of cardiovascular disease, systolic blood pressure, and total cholesterol. **Abbreviations:** eGFR: Estimated glomerular filtration rate.

| **Characteristic** | **HR**^1^ | **95% CI**^1^ | **p-value** |
| --- | --- | --- | --- |
| **Male sex** | 1.57 | 1.48, 1.67 | <0.001 |
| **Residence in Coyoacán** | 0.74 | 0.70, 0.78 | <0.001 |
| **Educational level** |  |  |  |
| University | Ref. | Ref. |  |
| High school | 1.16 | 1.02, 1.32 | 0.025 |
| Elementary | 1.39 | 1.24, 1.55 | <0.001 |
| Other | 1.46 | 1.29, 1.65 | <0.001 |
| **Smoking** |  |  |  |
| Never | Ref. | Ref. |  |
| Former | 1.14 | 1.07, 1.22 | <0.001 |
| Current | 1.15 | 1.07, 1.23 | <0.001 |
| **Diabetes** | 2.07 | 1.97, 2.19 | <0.001 |
| **Cardiovascular disease** | 1.67 | 1.53, 1.83 | <0.001 |
| **Systolic blood pressure (per mmHg)** | 1.00 | 1.00, 1.00 | <0.001 |
| **Total cholesterol (per mmol/L)** | 0.88 | 0.85, 0.90 | <0.001 |
| **eGFR estimated using the 2009 equation** |  |  |  |
| rcs(EFGR.09) | 0.97 | 0.97, 0.97 | <0.001 |
| rcs(EFGR.09)' | 1.03 | 1.02, 1.04 | <0.001 |
| rcs(EFGR.09)'' | 0.72 | 0.60, 0.86 | <0.001 |
| rcs(EFGR.09) ''' | 4.13 | 2.34, 7.31 | <0.001 |
| ^1^HR = Hazard Ratio, CI = Confidence Interval | | | |

**Supplementary Table 12.** Cox proportional hazard model for prediction of **all-cause mortality** at **5-years** using MCPS data and the **2021 CKD-EPI equation** for eGFR. The baseline model was stratified by age category and adjusted for sex, place of residence (Coyoacán or Iztapalapa), educational level (elementary, high school, university, other), smoking, diabetes, history of cardiovascular disease, systolic blood pressure, and total cholesterol. **Abbreviations:** eGFR: Estimated glomerular filtration rate.

| **Characteristic** | **HR**^1^ | **95% CI**^1^ | **p-value** |
| --- | --- | --- | --- |
| **Male sex** | 1.54 | 1.48, 1.60 | <0.001 |
| **Residence in Coyoacán** | 0.75 | 0.72, 0.78 | <0.001 |
| **Educational level** |  |  |  |
| University | Ref. | Ref. |  |
| High school | 1.18 | 1.08, 1.28 | <0.001 |
| Elementary | 1.39 | 1.29, 1.50 | <0.001 |
| Other | 1.44 | 1.33, 1.57 | <0.001 |
| **Smoking** |  |  |  |
| Never | Ref. | Ref. |  |
| Former | 1.08 | 1.03, 1.13 | 0.001 |
| Current | 1.12 | 1.07, 1.18 | <0.001 |
| **Diabetes** | 2.22 | 2.14, 2.30 | <0.001 |
| **Cardiovascular disease** | 1.52 | 1.42, 1.62 | <0.001 |
| **Systolic blood pressure (per mmHg)** | 1.00 | 1.00, 1.00 | <0.001 |
| **Total cholesterol (per mmol/L)** | 0.92 | 0.90, 0.93 | <0.001 |
| **eGFR estimated using the 2009 equation** |  |  |  |
| rcs(EFGR.09) | 0.98 | 0.98, 0.98 | <0.001 |
| rcs(EFGR.09)' | 1.02 | 1.01, 1.03 | <0.001 |
| rcs(EFGR.09)'' | 0.84 | 0.75, 0.94 | 0.002 |
| rcs(EFGR.09) ''' | 1.94 | 1.43, 2.63 | <0.001 |
| ^1^HR = Hazard Ratio, CI = Confidence Interval | | | |

**Supplementary Table 13.** Cox proportional hazard model for prediction of **all-cause mortality** at **10-years** using MCPS data and the **2009 CKD-EPI equation** for eGFR. The baseline model was stratified by age category and adjusted for sex, place of residence (Coyoacán or Iztapalapa), educational level (elementary, high school, university, other), smoking, diabetes, history of cardiovascular disease, systolic blood pressure, and total cholesterol. **Abbreviations:** eGFR: Estimated glomerular filtration rate.

| **Characteristic** | **HR**^1^ | **95% CI**^1^ | **p-value** |
| --- | --- | --- | --- |
| **Male sex** | 1.55 | 1.48, 1.61 | <0.001 |
| **Residence in Coyoacán** | 0.75 | 0.72, 0.78 | <0.001 |
| **Educational level** |  |  |  |
| University | Ref. | Ref. |  |
| High school | 1.18 | 1.08, 1.28 | <0.001 |
| Elementary | 1.39 | 1.29, 1.51 | <0.001 |
| Other | 1.44 | 1.33, 1.57 | <0.001 |
| **Smoking** |  |  |  |
| Never | Ref. | Ref. |  |
| Former | 1.08 | 1.03, 1.13 | 0.001 |
| Current | 1.12 | 1.07, 1.18 | <0.001 |
| **Diabetes** | 2.22 | 2.14, 2.30 | <0.001 |
| **Cardiovascular disease** | 1.52 | 1.42, 1.62 | <0.001 |
| **Systolic blood pressure (per mmHg)** | 1.00 | 1.00, 1.00 | <0.001 |
| **Total cholesterol (per mmol/L)** | 0.92 | 0.90, 0.93 | <0.001 |
| **eGFR estimated using the 2009 equation** |  |  |  |
| rcs(EFGR.09) | 0.98 | 0.98, 0.98 | <0.001 |
| rcs(EFGR.09)' | 1.02 | 1.01, 1.02 | <0.001 |
| rcs(EFGR.09)'' | 0.84 | 0.74, 0.95 | 0.004 |
| rcs(EFGR.09) ''' | 2.40 | 1.64, 3.51 | <0.001 |
| ^1^HR = Hazard Ratio, CI = Confidence Interval | | | |

**Supplementary Table 14.** Cox proportional hazard model for prediction of **all-cause mortality** at **10-years** using MCPS data and the **2021 CKD-EPI equation** for eGFR. The baseline model was stratified by age category and adjusted for sex, place of residence (Coyoacán or Iztapalapa), educational level (elementary, high school, university, other), smoking, diabetes, history of cardiovascular disease, systolic blood pressure, and total cholesterol. **Abbreviations:** eGFR: Estimated glomerular filtration rate.

| **Characteristic** | **sHR**^1^ | **95% CI**^1^ | **p-value** |
| --- | --- | --- | --- |
| **Male sex** | 1.46 | 1.31, 1.62 | <0.001 |
| **Residence in Coyoacán** | 0.75 | 0.68, 0.83 | <0.001 |
| **Educational level** |  |  |  |
| University | Ref. | Ref. |  |
| High school | 1.14 | 0.91, 1.43 | 0.3 |
| Elementary | 1.24 | 1.01, 1.51 | 0.037 |
| Other | 1.16 | 0.94, 1.44 | 0.2 |
| **Smoking** |  |  |  |
| Never | Ref. | Ref. |  |
| Former | 1.18 | 1.05, 1.32 | 0.005 |
| Current | 1.17 | 1.03, 1.33 | 0.017 |
| **Diabetes** | 1.87 | 1.71, 2.05 | <0.001 |
| **Cardiovascular disease** | 2.55 | 2.24, 2.89 | <0.001 |
| **Systolic blood pressure (per mmHg)** | 1.01 | 1.00, 1.01 | <0.001 |
| **Total cholesterol (per mmol/L)** | 0.92 | 0.88, 0.96 | <0.001 |
| **eGFR estimated using the 2009 equation** |  |  |  |
| rcs(EFGR.09) | 0.98 | 0.98, 0.98 | <0.001 |
| rcs(EFGR.09)' | 1.00 | 0.98, 1.02 | 0.8 |
| rcs(EFGR.09)'' | 0.91 | 0.66, 1.25 | 0.6 |
| rcs(EFGR.09) ''' | 2.00 | 0.75, 5.34 | 0.2 |
| ^1^sHR = subdistribution Hazard Ratio, CI = Confidence Interval | | | |

**Supplementary Table 15.** Fine & Gray model for prediction of **cardiovascular mortality** at **5-years** using MCPS data and the **2009 CKD-EPI equation** for eGFR. The baseline model was stratified by age category and adjusted for sex, place of residence (Coyoacán or Iztapalapa), educational level (elementary, high school, university, other), smoking, diabetes, history of cardiovascular disease, systolic blood pressure, and total cholesterol. **Abbreviations:** eGFR: Estimated glomerular filtration rate.

| **Characteristic** | **sHR**^1^ | **95% CI**^1^ | **p-value** |
| --- | --- | --- | --- |
| **Male sex** | 1.47 | 1.32, 1.63 | <0.001 |
| **Residence in Coyoacán** | 0.75 | 0.68, 0.83 | <0.001 |
| **Educational level** |  |  |  |
| University | Ref. | Ref. |  |
| High school | 1.14 | 0.91, 1.43 | 0.3 |
| Elementary | 1.24 | 1.01, 1.51 | 0.036 |
| Other | 1.16 | 0.94, 1.44 | 0.2 |
| **Smoking** |  |  |  |
| Never | Ref. | Ref. |  |
| Former | 1.18 | 1.05, 1.32 | 0.005 |
| Current | 1.17 | 1.03, 1.33 | 0.018 |
| **Diabetes** | 1.87 | 1.71, 2.05 | <0.001 |
| **Cardiovascular disease** | 2.55 | 2.24, 2.89 | <0.001 |
| **Systolic blood pressure (per mmHg)** | 1.01 | 1.00, 1.01 | <0.001 |
| **Total cholesterol (per mmol/L)** | 0.92 | 0.88, 0.96 | <0.001 |
| **eGFR estimated using the 2009 equation** |  |  |  |
| rcs(EFGR.09) | 0.98 | 0.98, 0.99 | <0.001 |
| rcs(EFGR.09)' | 0.99 | 0.97, 1.01 | 0.4 |
| rcs(EFGR.09)'' | 0.87 | 0.61, 1.25 | 0.5 |
| rcs(EFGR.09) ''' | 2.90 | 0.84, 10.0 | 0.093 |
| ^1^sHR = subdistribution Hazard Ratio, CI = Confidence Interval | | | |

**Supplementary Table 16.** Fine & Gray model for prediction of **cardiovascular mortality** at **5-years** using MCPS data and the **2021 CKD-EPI equation** for eGFR. The baseline model was stratified by age category and adjusted for sex, place of residence (Coyoacán or Iztapalapa), educational level (elementary, high school, university, other), smoking, diabetes, history of cardiovascular disease, systolic blood pressure, and total cholesterol. **Abbreviations:** eGFR: Estimated glomerular filtration rate.

| **Characteristic** | **HR**^1^ | **95% CI**^1^ | **p-value** |
| --- | --- | --- | --- |
| **Male sex** | 1.40 | 1.30, 1.51 | <0.001 |
| **Residence in Coyoacán** | 0.76 | 0.71, 0.81 | <0.001 |
| **Educational level** |  |  |  |
| University | Ref. | Ref. |  |
| High school | 1.15 | 0.99, 1.33 | 0.075 |
| Elementary | 1.20 | 1.05, 1.37 | 0.007 |
| Other | 1.16 | 1.01, 1.34 | 0.042 |
| **Smoking** |  |  |  |
| Never | Ref. | Ref. |  |
| Former | 1.12 | 1.03, 1.21 | 0.006 |
| Current | 1.12 | 1.03, 1.22 | 0.010 |
| **Diabetes** | 1.90 | 1.79, 2.02 | <0.001 |
| **Cardiovascular disease** | 2.24 | 2.03, 2.46 | <0.001 |
| **Systolic blood pressure (per mmHg)** | 1.01 | 1.01, 1.01 | <0.001 |
| **Total cholesterol (per mmol/L)** | 0.96 | 0.93, 0.99 | 0.008 |
| **eGFR estimated using the 2009 equation** |  |  |  |
| rcs(EFGR.09) | 0.99 | 0.99, 0.99 | <0.001 |
| rcs(EFGR.09)' | 0.99 | 0.97, 1.00 | 0.11 |
| rcs(EFGR.09)'' | 0.98 | 0.80, 1.21 | 0.9 |
| rcs(EFGR.09) ''' | 1.58 | 0.84, 2.94 | 0.2 |
| ^1^sHR = subdistribution Hazard Ratio, CI = Confidence Interval | | | |

**Supplementary Table 17.** Fine & Gray model for prediction of **cardiovascular mortality** at **10-years** using MCPS data and the **2009 CKD-EPI equation** for eGFR. The baseline model was stratified by age category and adjusted for sex, place of residence (Coyoacán or Iztapalapa), educational level (elementary, high school, university, other), smoking, diabetes, history of cardiovascular disease, systolic blood pressure, and total cholesterol. **Abbreviations:** eGFR: Estimated glomerular filtration rate.

| **Characteristic** | **HR**^1^ | **95% CI**^1^ | **p-value** |
| --- | --- | --- | --- |
| **Male sex** | 1.41 | 1.31, 1.51 | <0.001 |
| **Residence in Coyoacán** | 0.76 | 0.71, 0.81 | <0.001 |
| **Educational level** |  |  |  |
| University | Ref. | Ref. |  |
| High school | 1.15 | 0.99, 1.33 | 0.074 |
| Elementary | 1.20 | 1.05, 1.37 | 0.007 |
| Other | 1.16 | 1.01, 1.34 | 0.041 |
| **Smoking** |  |  |  |
| Never | Ref. | Ref. |  |
| Former | 1.12 | 1.03, 1.21 | 0.006 |
| Current | 1.12 | 1.03, 1.22 | 0.010 |
| **Diabetes** | 1.90 | 1.79, 2.03 | <0.001 |
| **Cardiovascular disease** | 2.24 | 2.03, 2.46 | <0.001 |
| **Systolic blood pressure (per mmHg)** | 1.01 | 1.01, 1.01 | <0.001 |
| **Total cholesterol (per mmol/L)** | 0.96 | 0.93, 0.99 | 0.007 |
| **eGFR estimated using the 2009 equation** |  |  |  |
| rcs(EFGR.09) | 0.99 | 0.99, 0.99 | <0.001 |
| rcs(EFGR.09)' | 0.99 | 0.97, 1.00 | 0.036 |
| rcs(EFGR.09)'' | 0.96 | 0.76, 1.21 | 0.7 |
| rcs(EFGR.09) ''' | 2.01 | 0.92, 4.42 | 0.081 |
| ^1^sHR = subdistribution Hazard Ratio, CI = Confidence Interval | | | |

**Supplementary Table 18.** Fine & Gray model for prediction of **cardiovascular mortality** at **10-years** using MCPS data and the **2021 CKD-EPI equation** for eGFR. The baseline model was stratified by age category and adjusted for sex, place of residence (Coyoacán or Iztapalapa), educational level (elementary, high school, university, other), smoking, diabetes, history of cardiovascular disease, systolic blood pressure, and total cholesterol. eGFR: Estimated glomerular filtration rate.

| **Characteristic** | **HR**^1^ | **95% CI**^1^ | **p-value** |
| --- | --- | --- | --- |
| **Male sex** | 1.62 | 1.37, 1.92 | <0.001 |
| **Residence in Coyoacán** | 0.64 | 0.55, 0.76 | <0.001 |
| **Educational level** |  |  |  |
| University | Ref. | Ref. |  |
| High school | 2.03 | 1.31, 3.15 | 0.002 |
| Elementary | 2.57 | 1.70, 3.87 | <0.001 |
| Other | 2.85 | 1.85, 4.39 | <0.001 |
| **Smoking** |  |  |  |
| Never | Ref. | Ref. |  |
| Former | 1.13 | 0.94, 1.36 | 0.2 |
| Current | 1.01 | 0.82, 1.24 | >0.9 |
| **Diabetes** | 6.51 | 5.43, 7.81 | <0.001 |
| **Cardiovascular disease** | 1.17 | 0.90, 1.52 | 0.2 |
| **Systolic blood pressure (per mmHg)** | 1.01 | 1.00, 1.01 | <0.001 |
| **Total cholesterol (per mmol/L)** | 1.04 | 0.98, 1.11 | 0.2 |
| **eGFR estimated using the 2009 equation** |  |  |  |
| rcs(EFGR.09) | 0.95 | 0.95, 0.95 | <0.001 |
| rcs(EFGR.09)' | 1.01 | 0.97, 1.05 | 0.7 |
| rcs(EFGR.09)'' | 0.85 | 0.48, 1.52 | 0.6 |
| rcs(EFGR.09) ''' | 2.43 | 0.43, 13.8 | 0.3 |
| ^1^sHR = subdistribution Hazard Ratio, CI = Confidence Interval | | | |

**Supplementary Table 19.** Fine & Gray model for prediction of **kidney-related mortality** at **5-years** using MCPS data and the **2009 CKD-EPI equation** for eGFR. The baseline model was stratified by age category and adjusted for sex, place of residence (Coyoacán or Iztapalapa), educational level (elementary, high school, university, other), smoking, diabetes, history of cardiovascular disease, systolic blood pressure, and total cholesterol. eGFR: Estimated glomerular filtration rate.

| **Characteristic** | **HR**^1^ | **95% CI**^1^ | **p-value** |
| --- | --- | --- | --- |
| **Male sex** | 1.64 | 1.39, 1.95 | <0.001 |
| **Residence in Coyoacán** | 0.64 | 0.55, 0.76 | <0.001 |
| **Educational level** |  |  |  |
| University | Ref. | Ref. |  |
| High school | 2.03 | 1.31, 3.15 | 0.002 |
| Elementary | 2.57 | 1.70, 3.87 | <0.001 |
| Other | 2.85 | 1.85, 4.39 | <0.001 |
| **Smoking** |  |  |  |
| Never | Ref. | Ref. |  |
| Former | 1.13 | 0.94, 1.36 | 0.2 |
| Current | 1.01 | 0.82, 1.24 | >0.9 |
| **Diabetes** | 6.50 | 5.42, 7.80 | <0.001 |
| **Cardiovascular disease** | 1.17 | 0.90, 1.52 | 0.2 |
| **Systolic blood pressure (per mmHg)** | 1.01 | 1.00, 1.01 | <0.001 |
| **Total cholesterol (per mmol/L)** | 1.04 | 0.98, 1.11 | 0.2 |
| **eGFR estimated using the 2009 equation** |  |  |  |
| rcs(EFGR.09) | 0.95 | 0.95, 0.96 | <0.001 |
| rcs(EFGR.09)' | 1.00 | 0.97, 1.04 | >0.9 |
| rcs(EFGR.09)'' | 0.83 | 0.44, 1.55 | 0.6 |
| rcs(EFGR.09) ''' | 3.08 | 0.35, 27.0 | 0.3 |
| ^1^sHR = subdistribution Hazard Ratio, CI = Confidence Interval | | | |

**Supplementary Table 20.** Fine & Gray model for prediction of **kidney-related mortality** at **5-years** using MCPS data and the **2021 CKD-EPI equation** for eGFR. The baseline model was stratified by age category and adjusted for sex, place of residence (Coyoacán or Iztapalapa), educational level (elementary, high school, university, other), smoking, diabetes, history of cardiovascular disease, systolic blood pressure, and total cholesterol. eGFR: Estimated glomerular filtration rate.

| **Characteristic** | **HR**^1^ | **95% CI**^1^ | **p-value** |
| --- | --- | --- | --- |
| **Male sex** | 1.71 | 1.52, 1.93 | <0.001 |
| **Residence in Coyoacán** | 0.69 | 0.62, 0.77 | <0.001 |
| **Educational level** |  |  |  |
| University | Ref. | Ref. |  |
| High school | 1.98 | 1.47, 2.67 | <0.001 |
| Elementary | 2.66 | 2.02, 3.50 | <0.001 |
| Other | 2.87 | 2.15, 3.85 | <0.001 |
| **Smoking** |  |  |  |
| Never | Ref. | Ref. |  |
| Former | 1.08 | 0.95, 1.23 | 0.2 |
| Current | 1.02 | 0.89, 1.17 | 0.8 |
| **Diabetes** | 7.96 | 7.03, 9.02 | <0.001 |
| **Cardiovascular disease** | 0.99 | 0.81, 1.22 | >0.9 |
| **Systolic blood pressure (per mmHg)** | 1.01 | 1.00, 1.01 | <0.001 |
| **Total cholesterol (per mmol/L)** | 1.03 | 0.98, 1.08 | 0.2 |
| **eGFR estimated using the 2009 equation** |  |  |  |
| rcs(EFGR.09) | 0.96 | 0.96, 0.96 | <0.001 |
| rcs(EFGR.09)' | 1.01 | 0.98, 1.03 | 0.7 |
| rcs(EFGR.09)'' | 1.00 | 0.72, 1.38 | >0.9 |
| rcs(EFGR.09) ''' | 1.52 | 0.60, 3.84 | 0.4 |
| ^1^sHR = subdistribution Hazard Ratio, CI = Confidence Interval | | | |

**Supplementary Table 21.** Fine & Gray model for prediction of **kidney-related mortality** at **10-years** using MCPS data and the **2009 CKD-EPI equation** for eGFR. The baseline model was stratified by age category and adjusted for sex, place of residence (Coyoacán or Iztapalapa), educational level (elementary, high school, university, other), smoking, diabetes, history of cardiovascular disease, systolic blood pressure, and total cholesterol. eGFR: Estimated glomerular filtration rate.

| **Characteristic** | **HR**^1^ | **95% CI**^1^ | **p-value** |
| --- | --- | --- | --- |
| **Male sex** | 1.73 | 1.53, 1.95 | <0.001 |
| **Residence in Coyoacán** | 0.69 | 0.62, 0.77 | <0.001 |
| **Educational level** |  |  |  |
| University | Ref. | Ref. |  |
| High school | 1.98 | 1.47, 2.67 | <0.001 |
| Elementary | 2.66 | 2.02, 3.50 | <0.001 |
| Other | 2.88 | 2.15, 3.85 | <0.001 |
| **Smoking** |  |  |  |
| Never | Ref. | Ref. |  |
| Former | 1.08 | 0.95, 1.23 | 0.2 |
| Current | 1.02 | 0.89, 1.17 | 0.8 |
| **Diabetes** | 7.96 | 7.03, 9.02 | <0.001 |
| **Cardiovascular disease** | 0.99 | 0.81, 1.22 | >0.9 |
| **Systolic blood pressure (per mmHg)** | 1.01 | 1.00, 1.01 | <0.001 |
| **Total cholesterol (per mmol/L)** | 1.03 | 0.98, 1.08 | 0.2 |
| **eGFR estimated using the 2009 equation** |  |  |  |
| rcs(EFGR.09) | 0.96 | 0.96, 0.96 | <0.001 |
| rcs(EFGR.09)' | 1.00 | 0.98, 1.02 | >0.9 |
| rcs(EFGR.09)'' | 1.01 | 0.71, 1.44 | >0.9 |
| rcs(EFGR.09) ''' | 1.71 | 0.54, 5.41 | 0.4 |
| ^1^sHR = subdistribution Hazard Ratio, CI = Confidence Interval | | | |

**Supplementary Table 22.** Fine & Gray model for prediction of **kidney-related mortality** at **10-years** using MCPS data and the **2021 CKD-EPI equation** for eGFR. The baseline model was stratified by age category and adjusted for sex, place of residence (Coyoacán or Iztapalapa), educational level (elementary, high school, university, other), smoking, diabetes, history of cardiovascular disease, systolic blood pressure, and total cholesterol. eGFR: Estimated glomerular filtration rate.
